# Impact of Inaccurately Labeled Data on the Performance of Multi-label Classification for Disease Recognition

**DOI:** 10.64898/2026.07.22.26358665

**Authors:** Sophie Schmiegel, Hannah Marchi, Marvin-Hendrik Röchter, Martin Rudwaleit, Christiane Fuchs

**Affiliations:** Faculty of Business Administration and Economics, Data Science Group, Bielefeld University, Germany; Hospital Bielefeld Rosenhöhe, Department of Internal Medicine and Rheumatology; Bielefeld University, Medical School and University Medical Center OWL; Institute of Computational Biology, Computational Health Center, Helmholtz Munich, Neuherberg, Germany

## Abstract

The process of medical diagnostics is challenging, especially since patients can simultaneously suffer from several diseases with similar, contradictory, or even opposing diagnoses. Statistical prediction can support physicians in this task; however, the quality of data used for predicition as well as the chosen statistical model can affect the reliability of data-driven decision support. Data quality can, in particular, be reduced by incomplete medical diagnoses, that is, the termination of the diagnostic process once a patient has tested positive for one disease that explains the symptoms. When interpreting missing diagnoses as negative, this leads to potentially false negative health data. Another source of low data quality lies in diagnoses being made through a principle of elimination, i.e., after several negative results, one opts for the seemingly last remaining possibility. This may lead to false positive health data.

In our work, we investigate how such inaccurately labeled data affects the predictive ability of multi-label classification (MLC) for disease recognition. Unlike single-label classification (SLC), MLC allows the simultaneous assignment of multiple diseases to a patient and can therefore describe clinical conditions more holistically.

To that end, we conduct a synthetic-data simulation study as well as a real-data case study on the example of chronic pain patients. In this regard, we compare MLC performance on accurately and inaccurately labeled data. We manipulate the data such that it corresponds to different diagnostic test sensitivities as well as to different examination sequences, thus paying special attention to resulting uncertainty within the process of medical diagnostics.

Our results show that inaccurate labeling substantially decreases MLC prediction ability. Furthermore, low diagnostic test-sensitivity, the order of disease examination and covariate effects have a strong impact on MLC performance. These findings contribute to a better understanding of the interplay and impact of diagnostic procedures, data documentation and interpretation, and statistical modeling. This underlines the need for careful data collection as a basis for model development; special consideration should be given to the extensive examination of patients as well as the targeted collection of covariates. This is particularly crucial when models are transferred into everyday clinical practice.

## 1 Introduction

The use of statistical models and artificial intelligence (AI) as basis of decision support systems is rapidly gaining importance. In the medical field, clinical decision support systems (CDSS) become more and more relevant (Sutton et al., 2020). Examples for CDSS are tools supporting physicians in drug prescriptions and the detection of drug-drug-interactions (e.g., Apidi et al., 2017), tools that help to assess visual symptoms (e.g., skin alterations) and other clinical findings in order to make accurate diagnoses (e.g., Barnett, 1987; Vardell and Bou-Crick, 2012; Martinez-Franco et al., 2018), and tools that provide compact information and latest updates about medical topics (e.g., Bradley-Ridout et al., 2021). In addition to these examples, efforts to enhance helpful tools are ongoing. Calcaterra et al. (2018), for example, developed a CDSS to assist physicians in appropriately deciding for or against carrying out image diagnostics. Further, Kunhimangalam et al. (2014) constructed a CDSS for diagnosing peripheral neuropathy. Their system is highly precise and achieves an accuracy of 93.3 %. The aforementioned examples and studies show that data-based approaches can significantly contribute to faster and more accurate diagnoses. However, they need to be based on valid and high-quality data to deliver reliable results as data quality substantially influences the performance of a CDSS (Sutton et al., 2020).

Technically, a CDSS is often based on a statistical model which is trained on data, collected for example in the context of medical examinations. Such data may be either prospective or retrospective in nature, which has important implications for their completeness and reliability (Bestehorn, 2014): Prospective data is collected for a predefined study purpose, ensuring that all relevant information is systematically considered and documented. In contrast, retrospective data is derived from existing health records, meaning that any inaccuracies, omissions, or incomplete information contained in those records are inevitably carried over into the dataset.

Diagnostic processes can be complex and may vary depending on the clinical setting, the professional experience of the examiner, and the patient’s condition. When symptoms cannot clearly be assigned to one single disease, patients are examined for several diseases with similar clinical symptoms, one of which may be a most suspected diagnosis and the other ones are so-called differential diagnoses. The physician decides on an examination sequence, that is the order in which a patient is tested for the various diseases. This decision is based on, among others, the physician’s suspicion, the duration of a test in combination with the urgency of treatment, the invasiveness of tests, the related cost, and resources available. In practice, several diagnostic tests may be run in parallel.

Whenever an suspected disease cannot be confirmed, the diagnostic process continues with the next disease until a positive diagnosis is made. At that point, taking the circumstances into account, the diagnostic process might be stopped. Hence, a patient is not necessarily examined for every differential diagnosis, avoiding overdiagnoses and psychological stress for the patient. However, the resulting health record neglects the fact that patients can simultaneously suffer from further diseases. In addition, a disease-by-disease examination that follows a principle of elimination may theoretically lead an a priori unlikely diagnosis, simply because none of the other diseases has been confirmed.

CDSS use the types of data described above as the basis for their predictions. In doing so, they transform health records into quantitative measures. From the diagnostic process just outlined, two potential sources of misinterpretation emerge, which may subsequently lead to errors in the data foundation of the CDSS: First, a disease that has not been examined is interpreted as being absent, although a patient may suffer from it; that is, a missing value is interpreted as ‘no’. Second, the final disease in an examination sequence is interpreted as being present due to the principle of elimination, although this is an incorrect diagnosis either; that is, a missing value is interpreted as ‘yes’. We refer to such data as inaccurately labeled data in the following.

In this work, we aim to investigate how such inaccurately labeled data affects the predictive ability of statistical classification for disease recognition. The statistical research question is motivated by the clinical need to effectively recognize diseases which cause chronic pain. In particular, we consider three such diseases: immune-mediated inflammatory diseases (IMID), the fibromyalgia syndrome (FMS), and osteoarthritis (OA). All of them cause chronic pain, and non-seldomly they occur simultaneously.

With respect to statistical classification, we distinguish between two approaches: single-label classification (SLC) and multi-label classification (MLC). These differ in terms of the number of classes (here: diseases) which a sample (here: patient) can be assigned to. SLC requires mutually exclusive classes, i.e., a sample is assigned to exactly one class, while MLC additionally allows the cases where a sample belongs to several, none or all classes. Because of the medical context outlined above, we focus on MLC in this work.

Although it may seem intuitive that poor training data quality leads to poor analysis results, it remains a research task to quantify this relationship precisely. Accordingly, this topic has been frequently discussed in the literature in various contexts. Feng et al. (2022), for example, highlight the need for a constant update of AI- and machine learning-based algorithms used in clinical practice. Getting there, the authors point out the need of high-quality training data to aim at reliable model updates. Schwabe et al. (2024) state that training data quality strongly affects the performance of deep learning models. Blake and Mangiameli (2011) investigate the impact of data quality on classification, assessing quality by four dimensions: accuracy, completeness, timeliness and consistency. They conclude that these factors influence the classification outcome, in addition to problem complexity. In our own previous work, we describe the process of developing a CDSS for antibiotic treatment in sepsis patients (Schmiegel et al., 2026a); our results underline the need for high-quality data as a basis of reliable tools for the use in clinical practice. Data quality comprises many aspects, including measurement error, measurement imprecision, misinterpretation, missing values, sparsity, missing meta-information, insufficient sample sizes, sampling bias, hidden confounders, and many others. The above overview highlights only a small subset of these factors. In this work, we focus on data quality in terms of the previously described incomplete diagnoses and the associated misinterpretations, in combination with the sensitivity of diagnostic tests, and we examine their impact on MLC. To the best of our knowledge, no prior study has addressed this issue.

In Section 2, we briefly introduce the statistical framework of MLC. In addition, we provide more detail on the medical context of chronic pain patients and the available data, together with a statistical model and assumptions on the data-generating process. In Section 3 we conduct a synthetic-data-based simulation study in which we investigate the impact of different kinds of data manipulation. The insights from the simulation study are further examined in a real-data-based case study in Section 4, where we estimate MLC models on both the original and on manipulated data. We report and discuss the results of the simulation study and case study in the respective sections, before Section 5 concludes the work and provides an outlook on future research.

## 2 Methods

### 2.1 Multi-label Classification for Disease Recognition

SLC and MLC methods are frequently used in medical diagnostics to classify patients (e.g., Farooq et al., 2017; Wosiak et al., 2018). The main difference between the two frameworks lies in the number *C* of simultaneously assumed classes: For SLC, the classes are mutually exclusive, while MLC accounts for co-assigned classes. We provide a detailed and graphically visualized description of SLC and MLC and the difference between these two approaches in Schmiegel et al. (2026b).

Both approaches share that they map observed covariates ***x***_*i*_ = (*x*_*i*1_, *x*_*i*2_, …, *x*_*ip*_) of sample *i* to target variables 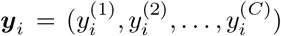. This mapping is learned on training data {***x***_*i*_, ***y***_*i*_} and subsequently applied to new data ***x***_*j*_, leading to predictions 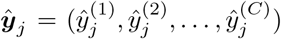 of the target variables. In this methodological introduction, we assume that ***y***_*i*_ is fully observed without error. The vectors ***y***_*i*_ and ***ŷ***_*i*_ are henceforth called true and predicted labelset, respectively; 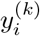 and 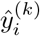 are called true and predicted labels for class *k* ∈ {1, …, *C* } . In our application, *i* ∈{1, …, *n* } indicates the *i*^*th*^ patient in a dataset, *n* is the total number of patients, *p* is the number of covariates, and *C* is the number of considered diseases. In SLC, one of the components of ***y***_*i*_ equals one, and all others equal zero; in MLC, each component of ***y***_*i*_ equals either zero or one without further restriction. We focus on MLC here.

To carry out MLC in practice, three workflows are distinguished, namely adaptation-based algorithms, ensemble-based methods or transformation-based methods (Herrera et al., 2016). Research gave rise to a large number of methods which can be assigned to these workflows (e.g., Sun et al., 2010; Bucak et al., 2011; Wu et al., 2016; Wehrmann et al., 2018; Xie and Huang, 2018). The three most prominent approaches belong to the group of transformation-based methods; these are binary relevance (Zhang et al., 2018), label powerset (Herrera et al., 2016) and classifier chains (CCs, Read et al., 2011). CCs predict the diseases under consideration in a stepwise manner, that is, the first examined disease is predicted using the observed covariates ***x***_*i*_; for the prediction of the second disease, both ***x***_*i*_ as well as the prediction 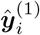 are used. This procedure is continued until the full labelset (i.e., all *C* labels) is predicted. In our work, we exclusively apply CCs because of three strengths: They can incorporate an association between disease occurrences, they can predict disease combinations even if they do not appear in the training data, and they can mirror the examination sequences in the estimation process.

As an essential component of CCs, a classifier needs to be chosen. We apply a logistic regression model (LRM); these models are well-established in various application fields including medicine, and they are computationally simple while showing a comparatively high predictive performance in other studies (e.g., Schmiegel et al., 2026b). For model optimization, we apply five-fold cross-validation (CV).

Common measures for prediction performance in the SLC context are sensitivity (not to be confused with the diagnostic test-sensitivity mentioned earlier), specificity, F1-score, precision and the area unter the curve (AUC). These measures can also be used in the MLC setting if one is interested in evaluating a prediction’s performance for each label separately. To assess the labels jointly, that is, as whole labelsets, the single measures are either averaged, or specific MLC measures are applied. Examples for the latter are the exact match ratio (EMR), Hamming loss and Jaccard similarity. EMR considers ***ŷ***_*i*_ to be correct if all components of ***ŷ***_*i*_ and ***y***_*i*_ match (e.g., Read et al., 2011) and indicates the fraction of correct predictions across all patients. EMR lies between zero (if there is no patient for which the labelset is correctly predicted) and one (if the labelset is correctly predicted for all patients). In contrast to this coarse but strict metric, the Jaccard similarity considers the number of true positive predictions divided by the sum of true positive, false positive and false negative predictions per patient, and then averages across patients (e.g., Bag et al., 2019). As EMR, the Jaccard similarity lies between zero and one, with value one indicating equality of both vectors throughout all patients. Last, the Hamming loss evaluates each label 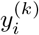 separately and computes the proportion of incorrectly predicted labels 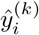 in the total number of labels across all patients (e.g., Read et al., 2011). As this metric is a loss function, a value close to zero is desirable.

Overall, we apply MLC, in particular CCs with LRM, and assess and compare the overall prediction performance by various established measures: To evaluate whole labelsets, we report EMR, Jaccard similarity as well as the Hamming loss; to evaluate individual labels, we regard sensitivity, specificity, precision, F1-score and AUC.

### 2.2 Clinical Use Case and Real Data Basis

Both our following simulation study and case study are based on data of chronic pain patients. Chronic pain is defined as persisting for more than three months or re-occurring within three months (Treede et al., 2015). The previously mentioned IMID, FMS, and OA belong to those diseases which cause chronic pain. In Germany, about 2.2 to 3.0 % of the adults suffer from IMID (Albrecht et al., 2023); further, the prevalence of FMS is 3.4 % (Häuser et al., 2020), while it is 17.9 % for OA (Fuchs et al., 2017). All of these diseases highly affect a patient’s quality of life (e.g., Russell et al., 2011). Assuming the diagnostic procedure being performed as described in Section 1, many occurrences might be missed. In case of IMID, for example, a lack of treatment can lead to long-term health consequences such as joint destruction (McInnes and Gravallese, 2021).

The considered data on chronic pain patients was collected by the Department of Internal Medicine and Rheumatology at Klinikum Bielefeld, Germany. At primary care, the patients were suspected to suffer from a rheumatic disease and were therefore referred to secondary care. None of the patients was pre-diagnosed with a rheumatic disease. The original dataset contained 152 covariates, collected at primary as well as at secondary care units, of 246 patients. Most covariates referred to various pain questionnaires, such as the ‘Fibromyalgia Rapid Screening Tool’ (FiRST; Perrot et al., 2010), the ‘Widespread Pain Index’ (Wolfe et al., 2010), the ‘Funktionsfragebogen Hannover’ (functional assessment questionnaire Hannover; Kohlmann and Raspe, 1996) and the ‘Central Sensitization Inventory’ (CSI; Mayer et al., 2011). The answers to individual questions of the questionnaires were included as separate covariates. Further, the dataset contained core data (e.g., age, sex, body mass index, professional background), measurements of C-reactive protein (CRP), information about the patients’ medical history as well as an initially suspected disease.

The data was collected as part of a prospective study. Therefore, all patients were carefully examined for all of the diseases of interest, that is, IMID, FMS and OA, independent of the suspected disease. Consequently, the underlying ground truth of every patient is known. For some patients, the data also contains information about the examiner’s a priori suspicion and further detected diseases, but this does not enter our analysis.

In Section 4, we describe the preprocessing of the real data in detail, which reduces the dataset to 164 patients. Here, we restrict ourselves to some key numbers: The prevalence of the three considered diseases IMID, FMS and OA are 29.9 %, 37.8 % and 28.0 %, respectively. Co-occurrences were observed in 27 out of 164 cases (16.5 %), whilst none of the three diseases was confirmed in 35 cases (21.3 %). The observed disease combinations are visualized in Figure 1. Figure A1 in the Appendix displays Cramér’s V, which quantifies the association between nominal variables, for the diseases. Accordingly, the association between IMID and FMS is moderate, while OA is hardly associated with any of the other two. While Section 4.1 describes all relevant covariates, five of them had already served as a suitable basis for synthetic data generation in a previous study (Marchi et al., 2026) and are also used accordingly in Section 3. These are CRP, age, sex, FiRST and numeric rating scale (NRS). The tender points, which are used as sixth covariate by Marchi et al. (2026), is replaced by CSI in our work since the tender points show a large number of missing values (see Figure A2 in the Appendix).

**Figure 1:**
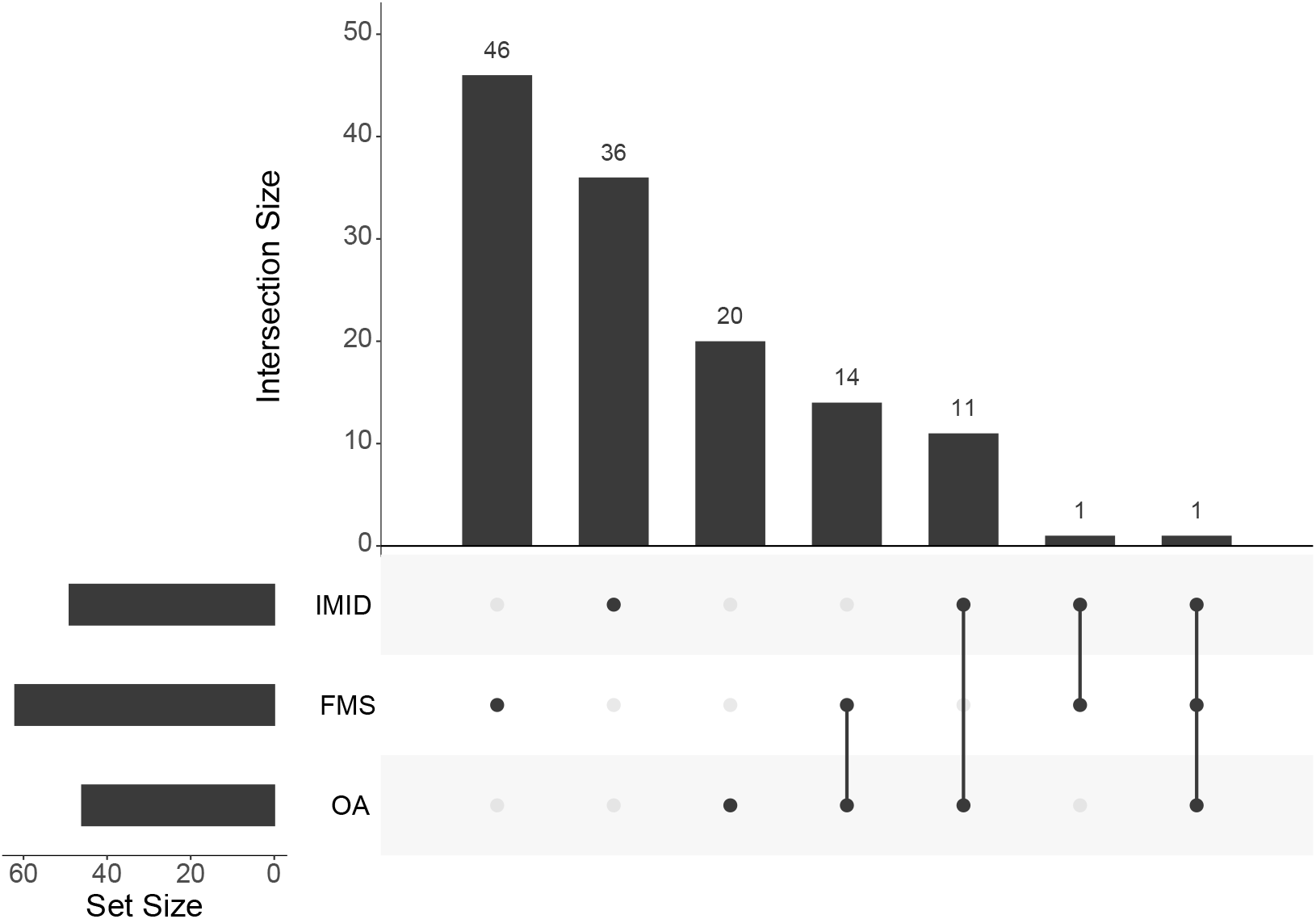
Visualization of the number of occurrences for each disease and their respective intersections in the final real dataset. A tabular representation of the numbers is provided in Table A1 in the Appendix. Accordingly, 35 patients do not suffer from any of the three diseases.

### 2.3 From Ground Truth to Interpreted Labelsets

We focus on the impact of inaccurately labeled data as it arises from a diagnostic process as described in Section 1. In the following, we formalize this process (making simplifying assumptions compared to clinical practice) and illustrate it using the concrete example of pain patients. Based on this formalization, we derive a data manipulation procedure that reflects such inaccurate labeling, which is subsequently applied in the simulation study in Section 3 and in the case study in Section 4.

#### 2.3.1 Inaccurate Labeling

Formally, we consider a patient *i* for whom *C* different diseases are considered possible based on the presenting symptoms. The true health state is coded through 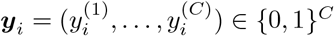, with the value one representing the presence of the respective disease. In particular, several diseases can apply simultaneously. This ground truth is unknown in practice. ***y***_*i*_ is called *true labelset*.

The examining physician determines an examination sequence consisting of *C* corresponding diagnostic tests. We assume that these tests are performed sequentially. If a test yields a positive result, the diagnostic process is terminated, as a cause of the symptoms has been identified. Otherwise, the sequence proceeds to the next test, up to the (*C*− 1)^th^ one. After that test, the diagnostic process is stopped in any case, because the first *C* − 1 test results already determine the *C*^th^ diagnosis. A truthful documentation of the test results yields a vector whose elements can take the values 0, 1, and NA (‘not available’), where NA marks tests that were not performed. We denote the resulting vector by ***r***_*i*_ and call it *recorded labelset*.

If a classifier is trained on labelsets, it typically cannot handle missing values directly. Therefore, NAs are usually replaced by zeros or ones. We assume that NAs are generally interpreted as zeros, with one exception: If the NA corresponds to the last test in the examination sequence and all other tests were negative, the result is set to one, reflecting a diagnosis by exclusion. We denote the resulting vector by ***z***_*i*_ and refer to it as the *interpreted labelset*.

A classifier is trained on such interpreted labelsets ***z***_*i*_, leading to *predicted labelsets* ***ŷ***_*i*_ by applying a learned mapping to covariates. The goal of a real application is for ***ŷ***_*i*_ and ***y***_*i*_ to agree as closely as possible. However, this is complicated by the fact that ***y***_*i*_ and ***z***_*i*_ may differ, and ***y***_*i*_ itself is unobserved. In our simulation study and case study, however, both are known, allowing us to quantify the impact of inaccurate labeling. The different kinds of labelsets are summarized in Table 1.

**Table 1:**
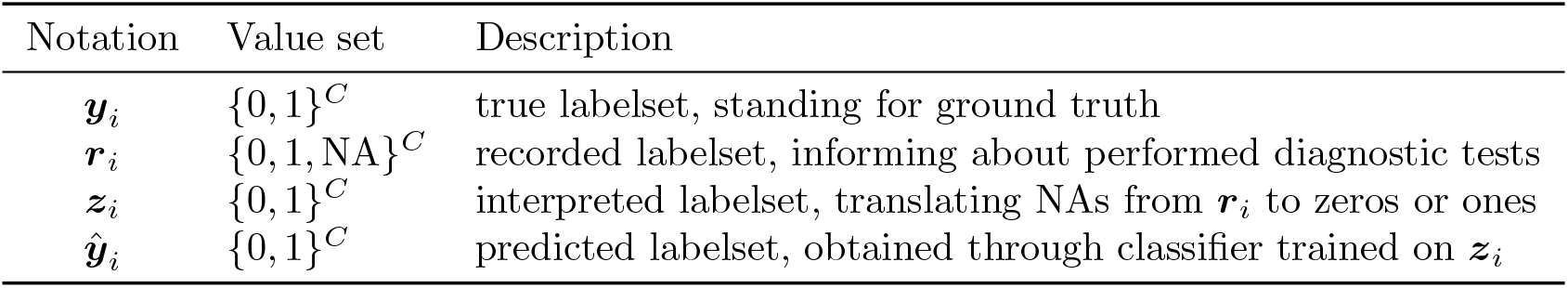
Kinds of labelsets considered in this work. We differentiate between the *true labelset* ***y***_*i*_, the *recorded labelset* ***r***_*i*_, the *interpreted labelset* ***z***_*i*_, and the *predicted labelset* ***ŷ***_*i*_ for a patient *i*.

| Notation | Value set | Description |
| --- | --- | --- |
| $\mathbf{y}_i$ | $\{0, 1\}^C$ | true labelset, standing for ground truth |
| $\mathbf{r}_i$ | $\{0, 1, \text{NA}\}^C$ | recorded labelset, informing about performed diagnostic tests |
| $\mathbf{z}_i$ | $\{0, 1\}^C$ | interpreted labelset, translating NAs from $\mathbf{r}_i$ to zeros or ones |
| $\hat{\mathbf{y}}_i$ | $\{0, 1\}^C$ | predicted labelset, obtained through classifier trained on $\mathbf{z}_i$ |

To further illustrate the diagnostic process and the resulting labelsets, we return to the context of chronic pain patients and the three possible diseases IMID, FMS and OA. Table 2 shows exemplary cases for possible ground truths (true labelsets) and examination sequences and the resulting recorded and interpreted labelsets. In Cases 1 to 3, we consider a hypothetical patient who suffers from FMS and OA, as can be seen from the true labels. The three cases differ in their examination sequence, which is represented by the order in which the respective diseases are listed. In *Case 1*, the patient is initially examined for IMID, but the test is negative. Next, the patient is tested for FMS, with a positive outcome, and therefore not tested for OA at all. The reported labels correctly represent these test results, but the previously described data interpretation leads to a translation of NAs into zeros or ones. In Case 1, this leads to the interpreted label of OA being absent, while the true label says that OA is actually present. A similar situation arises in *Case 2*, where FMS is tested first so that the diagnostic process is stopped afterwards. In *Case 3*, the reported labels are interpreted as OA being present while IMID and FMS are not. *Case 4* represents a patient who does not suffer from any of the considered diseases, but the assumed testing and interpretation mechanism labels IMID to be present following the principle of elimination. All in all, none of the listed cases leads to interpreted labelsets that entirely agree with the true labelsets. Our study investigates the impact of precisely this kind of inaccurate labeling on MLC performance.

**Table 2:** Examples of inaccurate labeling: Four cases of true labelsets and examination sequences, together with their diagnostic test results, leading to differing recorded and interpreted labelsets according to the assumed testing and interpretation process.

|  | True label | Examination sequence | Test result | Recorded label | Interpreted label |
| --- | --- | --- | --- | --- | --- |
| *Case 1* | 0 | IMID | negative | 0 | 0 |
|  | 1 | FMS | positive | 1 | 1 |
|  | 1 | OA | - | NA | 0 |
| *Case 2* | 1 | FMS | positive | 1 | 1 |
|  | 0 | IMID | - | NA | 0 |
|  | 1 | OA | - | NA | 0 |
| *Case 3* | 1 | OA | positive | 1 | 1 |
|  | 0 | IMID | - | NA | 0 |
|  | 1 | FMS | - | NA | 0 |
| *Case 4* | 0 | OA | negative | 0 | 0 |
|  | 0 | FMS | negative | 0 | 0 |
|  | 0 | IMID | - | 1 | 1 |

#### 2.3.2 Data Manipulation

To examine the impact of inaccurately labeled data on the predictive performance of MLC, we compare the performance of models estimated on accurately labeled data with models estimated on manipulated and thus inaccurately labeled data. In clinical practice, true labelsets are usually not available. In our studies they are: in the simulation study, because the ground truth is synthetically generated; and in the case study, because the data stems from a prospective patient cohort. Sections 3 and 4 provide details on that. In the following, we describe how we manipulate this data to mimic inaccurate labeling.

An important and so far neglected aspect of diagnostic processes is that diagnostic tests may be wrong, i.e., they can produce false-positive or false-negative results. This introduces an additional layer of uncertainty in the transition from true labelsets to recorded and subsequently interpreted labelsets. We account for the possible occurrence of false-negatives by introducing a diagnostic test-sensitivity *p*^(*k*)^ for disease *k* ∈ {1, …, *C*}. This parameter represents the probability that disease *k*, if truly present, is correctly detected by a positive test result. We restrict our analysis to incorporating diagnostic test-sensitivity rather than specificity, motivated by the example of chronic pain patients, where recognizing IMID is crucial to avoid long-term health consequences. In our studies, we consider the following scenarios for diagnostic test-sensitivities *p*^(1)^, …, *p*^(*C*)^: First, all diagnostic tests provide the truth with a test-sensitivity of 100 %; second, all diagnostic tests provide the truth with a test-sensitivity of 70 %; and third, the test-sensitivity varies across diseases, namely 85 % for disease A, 65 % for B and 100 % for C in the simulation study, and the same values for IMID, FMS and OA in the case study. The latter choice is motivated by clinical practice, where FMS is most difficult to diagnose and OA simplest. In Sections 3 and 4, the above procedure leads to MLC being performed on four kinds of datasets: the group TL of original true labelsets, and the groups IL_100_, IL_70_ and IL_diff_ of interpreted labelsets resulting from the just described scenarios of diagnostic test-sensitivity.

In both of our studies, we consider the case of three co-occurring diseases. We therefore describe the data manipulation process for *C* = 3 and illustrate it in Figure 2. Thus, consider a fictional or real patient *i* for which a true labelset 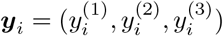 is available. Do the following:

1. Determine the sequence in which the diseases are examined. For the sake of simpler notation, we assume the sequence 1—2—3 in the following, but the process can be readily adapted.
2. If the patient suffers from the first examined disease, this will be recognized with the predefined test-sensitivity. Thus, if 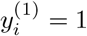, set 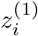 to one with probability *p*^(1)^. Otherwise, set 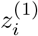 to zero.
3. If 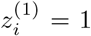, the diagnostic process stops and all remaining diseases are interpreted as being absent. In that case, set 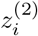 and 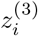 to zero.
4. Otherwise, continue with the second disease of the examination sequence: If 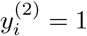, set 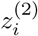 to one with probability *p*^(2)^. Otherwise, set 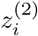 to zero.
5. The third disease will not be tested. If 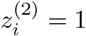, it is interpreted absent; in that case, set 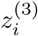 to zero. Otherwise, it is assumed to apply by exclusion of the other two; in that case, set 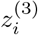 to one.

**Figure 2:**
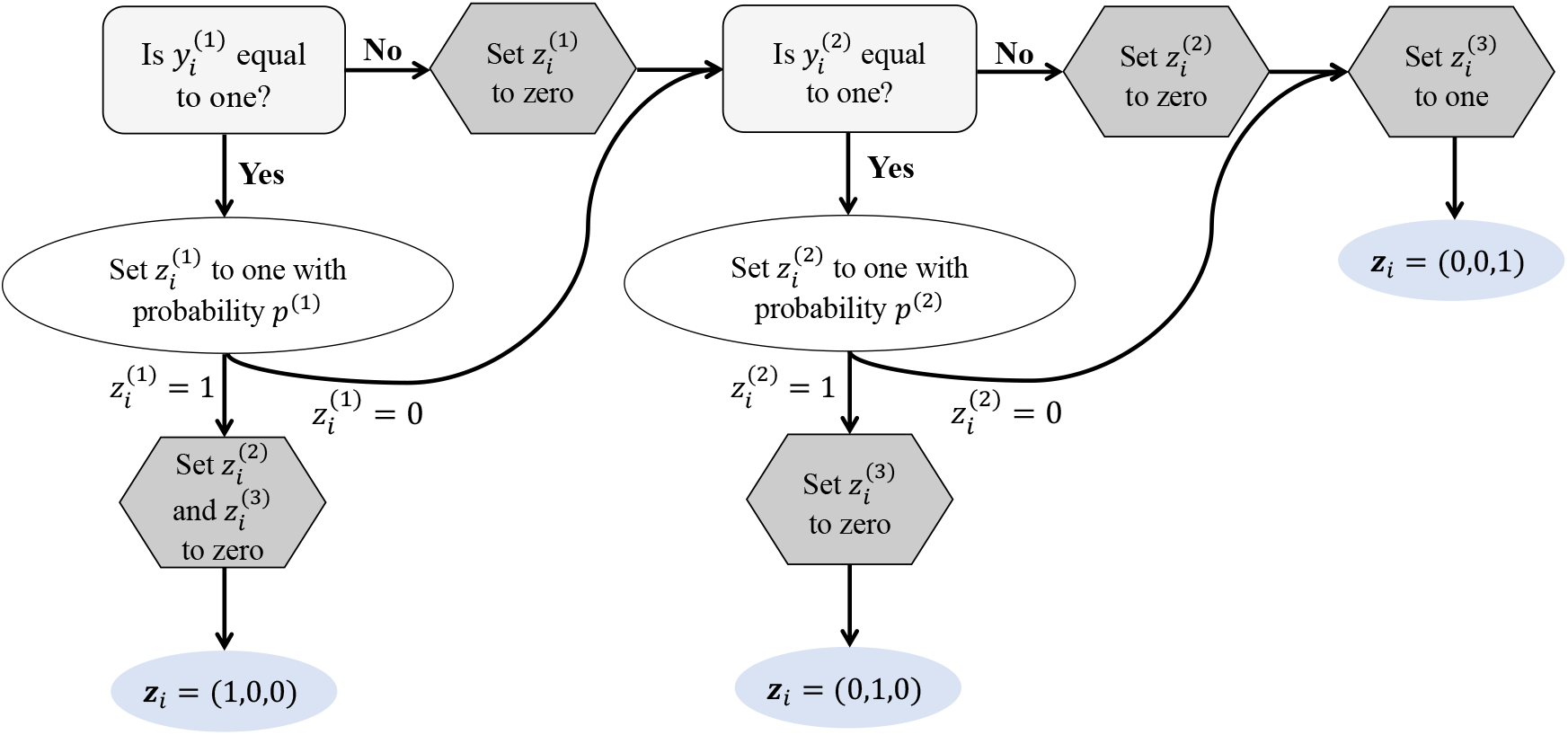
Flowchart of data manipulation from true labelset ***y***_*i*_ to interpreted labelset ***z***_*i*_ for simulation and case study.

## 3 Simulation Study

We conduct a simulation study to empirically evaluate the extent to which inaccurately labeled data affects the predictive performance of MLC. To that end, we generate synthetic data of multi-label type, such that the ground truth is known and can be compared against the prediction. We manipulate the data as described in Section 2.3.2 to assess the effect of inaccurate labeling. In particular, we train the classifier on four groups of datasets: TL, which contains the true labelsets; IL_100_, where inaccurate labeling occurs due to early termination of the diagnostic process and misinterpretation of missing values; IL_70_, which additionally imposes a diagnostic test-sensitivity of only 70 % for all diseases; and IL_diff_, where the test-sensitivity differs between diseases as specified in Section 2.3.2.

All estimated models are tested on accurately labeled data and compared regarding their ability to predict individual diseases (measured by sensitivity, specificity, precision, F1-score and AUC) and complete labelsets (measured by EMR, Jaccard similarity and Hamming loss). We further consider all possible examination sequences for each scenario.

In the following sections, we explain the process of data generation, describe the data resulting from this, and report the results which are subsequently discussed.

### 3.1 Synthetic Data Generation

While Section 2.3.2 described the process of manipulating the true labelsets to mimic diagnostic processes, errors and misinterpretation, it remains to be specified how the true labelsets are generated in the first place; in particular, in a way that induces both the desired associations between the covariates (i.e., patient characteristics) and the diseases, as well as dependencies among the diseases themselves. In the following, we generate synthetic true labelsets which are aligned with the presented context and the data situation of chronic pain patients. In particular, we consider three diseases (‘A’, ‘B’ and ‘C’) which we assume to be associated with each other, and establish an association to patient covariate data. The procedure of data generation follows the algorithmic framework proposed by Marchi et al. (2026).

To start with the patient covariate data, we consider six covariates *x*_1_, …, *x*_6_ of *n* fictional patients. As outlined in Section 2.2, we derive their characteristics from real-data information about CRP, age, sex, FiRST, NRS and CSI. Using the NORmal To Anything technique (Cario and Nelson, 1997), we generate fully synthetic covariate data, meaning that we derive patient characteristics from real health records, but other than that, real data points are ultimately excluded (McLachlan et al., 2019).

The covariates are subsequently used to simulate the target variables (disease occurrences) ***y***_*i*_ in a way such that *x*_1_, …, *x*_6_ have a predefined effect on ***y***_***i***_ and such that 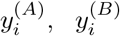 and 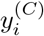 are associated. This is implemented by defining a logistic regression model for the effect of the covariates on the disease probabilities and subsequently simulating from this model. In particular, we introduce two standard normally distributed variables *U*_*AB*_ and *U*_*BC*_ which represent latent factors impacting the joint occurrences of diseases A and B and of diseases B and C, retrospectively. Both the covariates and the latent factors are used to simulate disease occurrences through a latent factor model as described as ‘Path C’ in Marchi et al. (2026).

To investigate the impact of the strength of covariate effects in the context of our research question, we define two sets of coefficients, henceforth called *Coefficient Set 1* and *Coefficient Set 2*. For *Coefficient Set 1*, we derive the coefficients from the real dataset; for *Coefficient Set 2*, we assume that each covariate influences the probability of occurrence for only two of the three diseases and thus modify the first coefficient set forcing selected coefficients to zero and then adapting the remaining coefficients to induce the associations displayed in Figure A3 in the Appendix. The corresponding regression coefficients are provided in Table A2 in the Appendix. The use of latent factors implies, however, that re-estimation of the regression coefficients will yield estimates deviating from the predefined values. For both coefficient sets, we choose the coefficients for the two latent variables to be 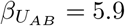 and 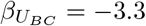.

We do not explicitly control for a dependence between occurrences of the three diseases. Instead, we induce different association strengths through covariate effects. In addition, we require marginal target prevalences of 45 % for disease A, 15 % for B and 60 % for C. Marchi et al. (2026) describe the underlying technique.

To further investigate the impact of sample size, we simulate training datasets of different numbers of patients, namely *n* ∈ {200, 1000 }. Together with the two described sets of coefficients, this results in four combinations of study settings. To additionally account for variation across individually generated datasets, we consider 500 independent replications of the procedure described below. In Section 3.2, we report results across these 500 replicates.

To attribute differences in the results specifically to the sources of interest in our study, we proceed as follows in the data simulation: For each replication, we generate patient covariate data ***x***_*i*_ for a large number of fictional patients (here: more than 1500), which constitute the study population for all sample sizes and coefficient sets. For each coefficient set, we simulate the true labelsets ***y***_*i*_ as described before. Afterwards, we separate 500 samples as test patients; the corresponding true labelsets are henceforth called ‘TL test sets’. From the remaining samples, we randomly select 1000 training patients, from which we again sample 200 patients; the corresponding true labelsets constitute the ‘TL training sets’. Before extracting the 200 or 1000 patients, we manipulate the target variables at the level of the full study population as described in Section 2.3.2. Consequently, the resulting inaccurately labeled datasets for the 200 patients correspond exactly to their respective entries in the larger dataset of size 1000.

To obtain the inaccurately labeled datasets (‘IL training sets’), we iterate through each of the six possible examination sequences (of the three diseases) and each of the considered diagnostic test-sensitivity settings. This results in 24 IL training sets per coefficient set, namely six each for IL_100_, IL_70_ and IL_diff_.

Table 3 displays characteristics of the covariate data across all replicates, excluding test patients. Cramér’s V for the three diseases (according to TL training sets) is visualized in Figure A3 in the Appendix for *n* = 200 and *n* = 1000 as well as for Coefficient Sets 1 and 2, averaged across the 500 replicates. Accordingly, in case of Coefficient Set 1, the diseases A and B as well as B and C are moderately associated, while A and C are hardly associated; in case of Coefficient Set 2, A and B are moderately associated, but C is hardly associated with any of the two diseases. Across all coefficient sets, sample sizes and replicates, this ultimately results in prevalence ranges of 44.4 % to 45.1 % for disease A, 14.1 % to 15.1 % for B and 59.9 % to 60.2 % for C. The empirically obtained prevalences do almost fully correspond to the predefined target prevalences.

**Table 3:**
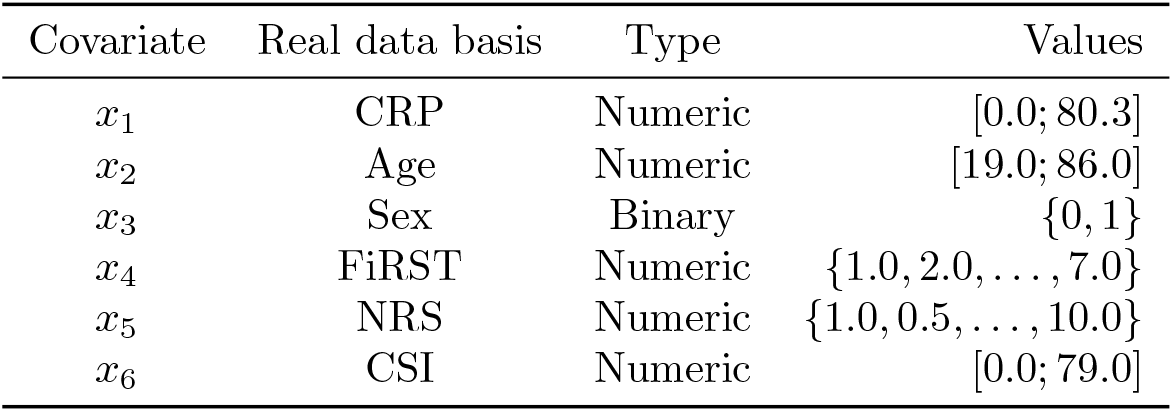
Characteristics of covariates used for simulation study. The values refer to the simulated study population across the 500 replicates.

| Covariate | Real data basis | Type | Values |
| --- | --- | --- | --- |
| $x_1$ | CRP | Numeric | [0.0; 80.3] |
| $x_2$ | Age | Numeric | [19.0; 86.0] |
| $x_3$ | Sex | Binary | {0, 1} |
| $x_4$ | FiRST | Numeric | {1.0, 2.0, ..., 7.0} |
| $x_5$ | NRS | Numeric | {1.0, 0.5, ..., 10.0} |
| $x_6$ | CSI | Numeric | [0.0; 79.0] |

### 3.2 Results

We estimate MLC models based on accurately as well as on inaccurately labeled data for the four combinations of training sample sizes and coefficient sets. Within each of these combinations, we consider all six examination sequences, which affect the process in two ways: On the one hand, the examination sequence is part of the manipulation of the true labelsets, as described in Section 2.3.2, and potentially leads to different interpreted labelsets; we train the classifier on each of the six versions of IL_100_, IL_70_ and IL_diff_. Second, the training of CCs employs a label ordering for which we always use the examination sequences corresponding to the IL training sets. Even for training on TL training sets, the examination sequence plays a role: While the underlying dataset remains unchanged, the CC label ordering is affected.

After training has been completed, the learned classifiers are applied to generate predictions for the test patients. These predictions are consistently evaluated against the same TL test sets for the respective coefficient set, regardless of the underlying examination sequence. Results are reported across 500 replicates. The model names and the specified sequences of diseases refer to the study settings described above.

#### Prediction of Full Labelsets

Figures 3 and 4 visualize measures for the prediction of full labelsets for Coefficient Sets 1 and 2, respectively. We report EMR and Jaccard similarity (where higher values indicate better predictive performance) here and include the Hamming loss in the Appendix (see Figures A4 and A5). The heights of the bars indicate average values across 500 replicates; black lines show their ranges. The colors distinguish between TL, IL_100_, IL_70_ and IL_diff_ models.

**Figure 3:**
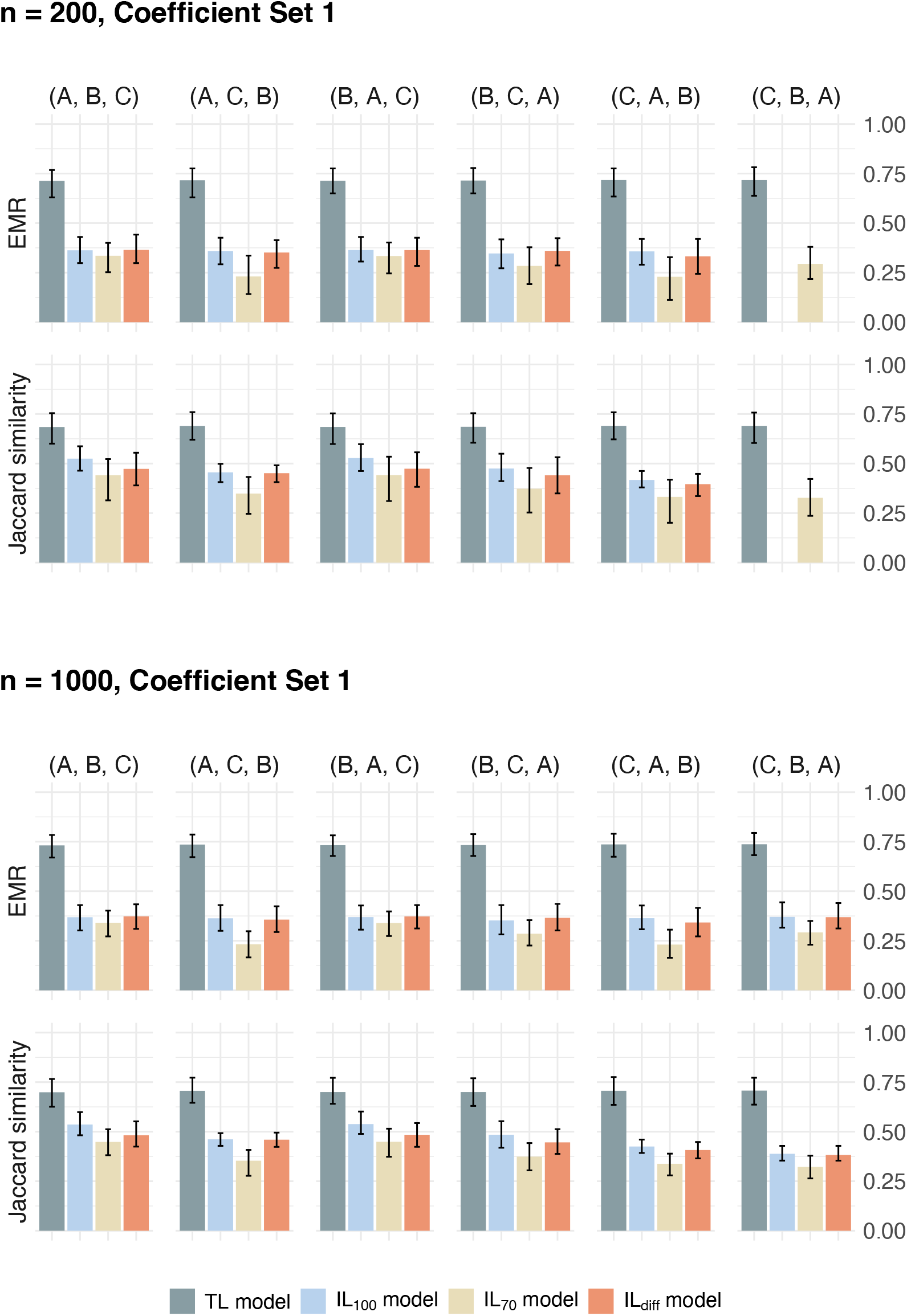
EMR and Jaccard similarity for the models trained on accurately and on inaccurately labeled synthetic data (different colored bars) considering Coefficient Set 1 for data generation and assuming different examination sequences (groups of four bars each). The model names refer to the employed training datasets. Higher values speak in favor of better model performance. Missing results stem from the fact that in some replicates, one of the diseases was only scarcely represented. This made the estimation impossible. Hamming loss is visualized in Figure A4 in the Appendix.

**Figure 4:**
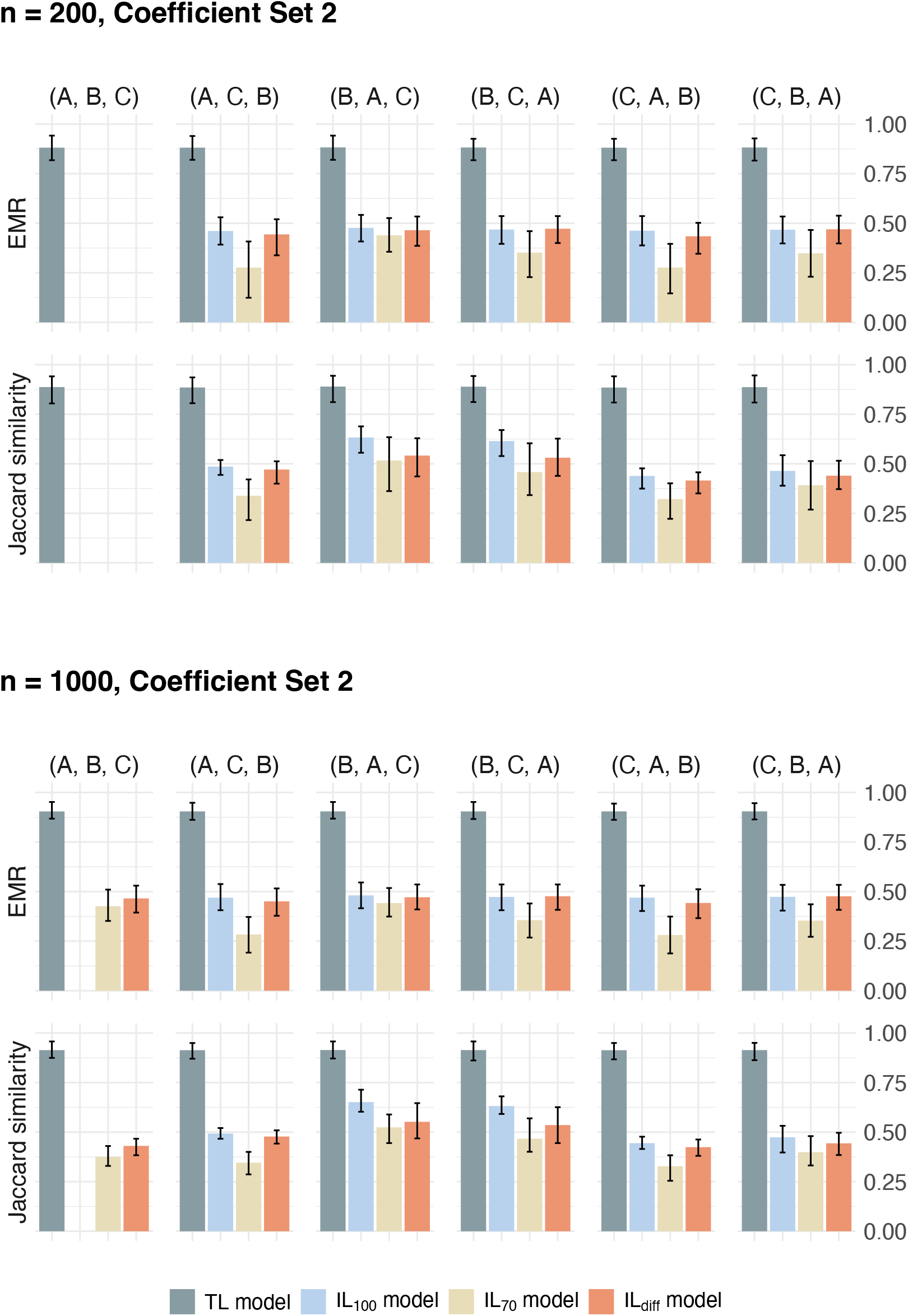
EMR and Jaccard similarity as in Figure 3, here for Coefficient Set 2. Hamming loss is visualized in Figure A5 in the Appendix.

TL models (gray-blue bars) clearly outperform all other models with respect to all displayed measures and all examination sequences. The difference between the three IL models, in contrast, is less clear: Looking at averages, the IL_100_ models (light blue bars) perform similar to the IL_diff_ models (orange bars) with respect to EMR, and in most cases slightly better regarding Jaccard similarity. The IL_70_ models (yellow bars) perform worst for both EMR and Jaccard similarity and all examination sequences. The same holds for Hamming loss (see Figures A4 and A5 in the Appendix).

A comparison of ranges across replicates, i.e. lowest and highest values achieved in one of the 500 replicates (indicated by the black lines), confirms the dominance of the TL models, for which the intervals never overlap with those of the IL models. Ranges are of comparable size across TL, IL_100_ and IL_diff_ models, and smaller than those of the IL_70_ models, which indicate particularly high variation for the Hamming loss.

Regarding sample size (i.e., comparing the upper and lower parts of Figures 3 and 4), average performances are comparable, but ranges decrease with increasing number of patients. Especially for the smaller sample size, no results could be obtained from some of the IL models as in some replicates, one of the diseases was only scarcely represented. This made the estimation of CCs on CV-folds partly impossible, meaning that the model estimation failed. Our observations also hold for the Hamming loss in Figures A4 and A5 in the Appendix.

When comparing the achieved performance based on the two coefficient sets, a markedly better performance of the TL models under Coefficient Set 2 becomes apparent: While the EMR across all sample sizes, examination sequences and replicates never exceeds 0.795 for Coefficient Set 1 and the Jaccard similarity never exceeds 0.776, the EMR for Coefficient Set 2 is consistently above 0.816 and the Jaccard similarity consistently above 0.804. For the IL models, the average performance under Coefficient Set 2 is often higher as well, but with exceptions.

#### Prediction of Individual Labels

Figures 5 and 6 visualize the ability of the models to predict individual diseases, that means: While each MLC model yields a three-dimensional prediction 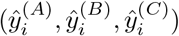, we evaluate this vector label-by-label. The colored cells of the heatmaps indicate the sensitivity, specificity, precision, F1-score and AUC for all three diseases A, B and C (labels on the left), every examination sequence (labels on the right) and each of the four TL and IL models (labels at the top). Dark colors depict better performance and, vice versa, light colors lower performance.

**Figure 5:**
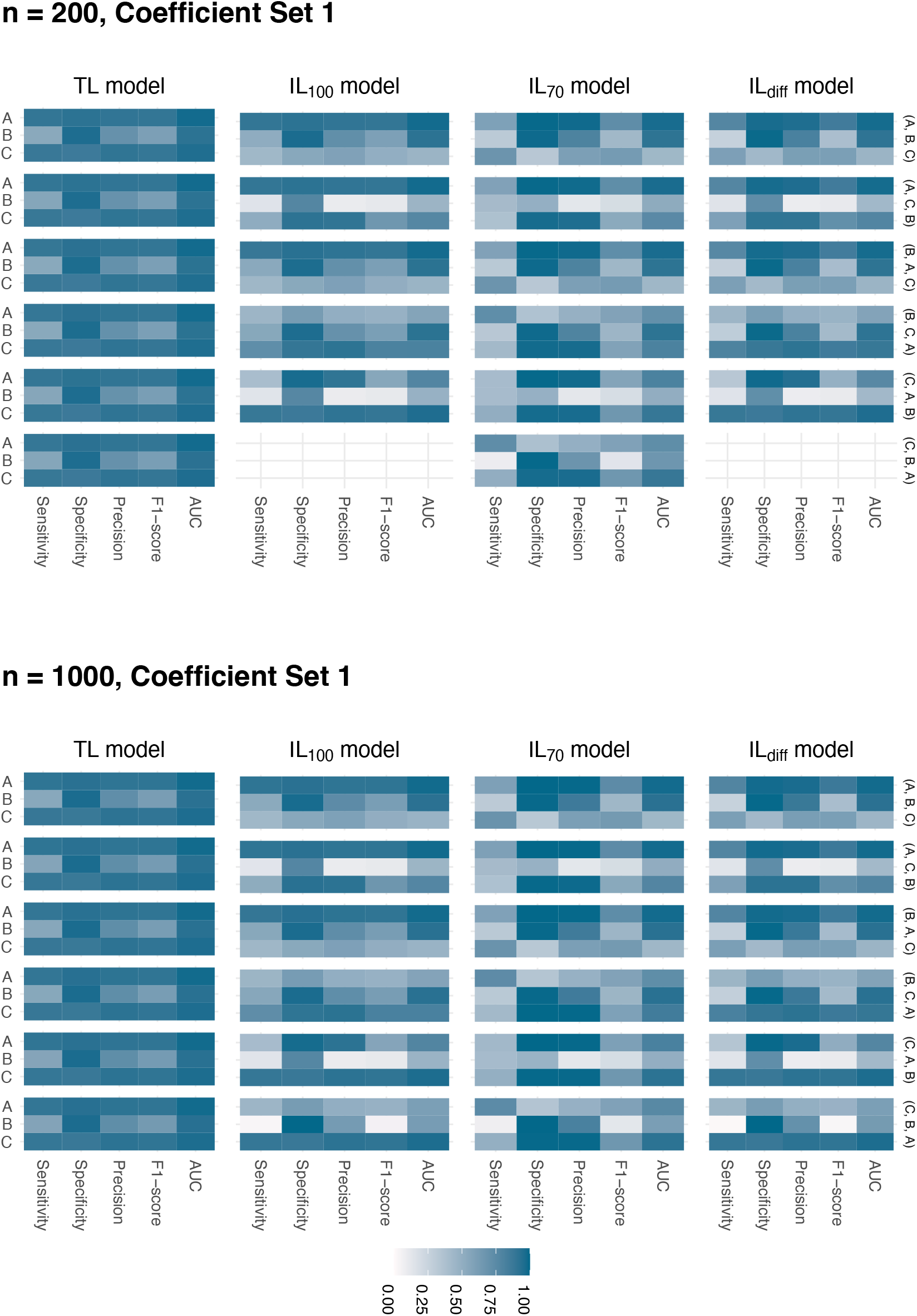
Heatmaps of performance of predicting individual diseases A, B and C (see labels on the left) for all models (labels at the top) and examination sequences (labels on the right) averaged across 500 replicates of size *n* = 200 (upper panel) and *n* = 1000 (bottom panel), using Coefficient Set 1 for data generation. Dark colors depict larger values and vice versa. The model names refer to the employed training datasets.

**Figure 6:**
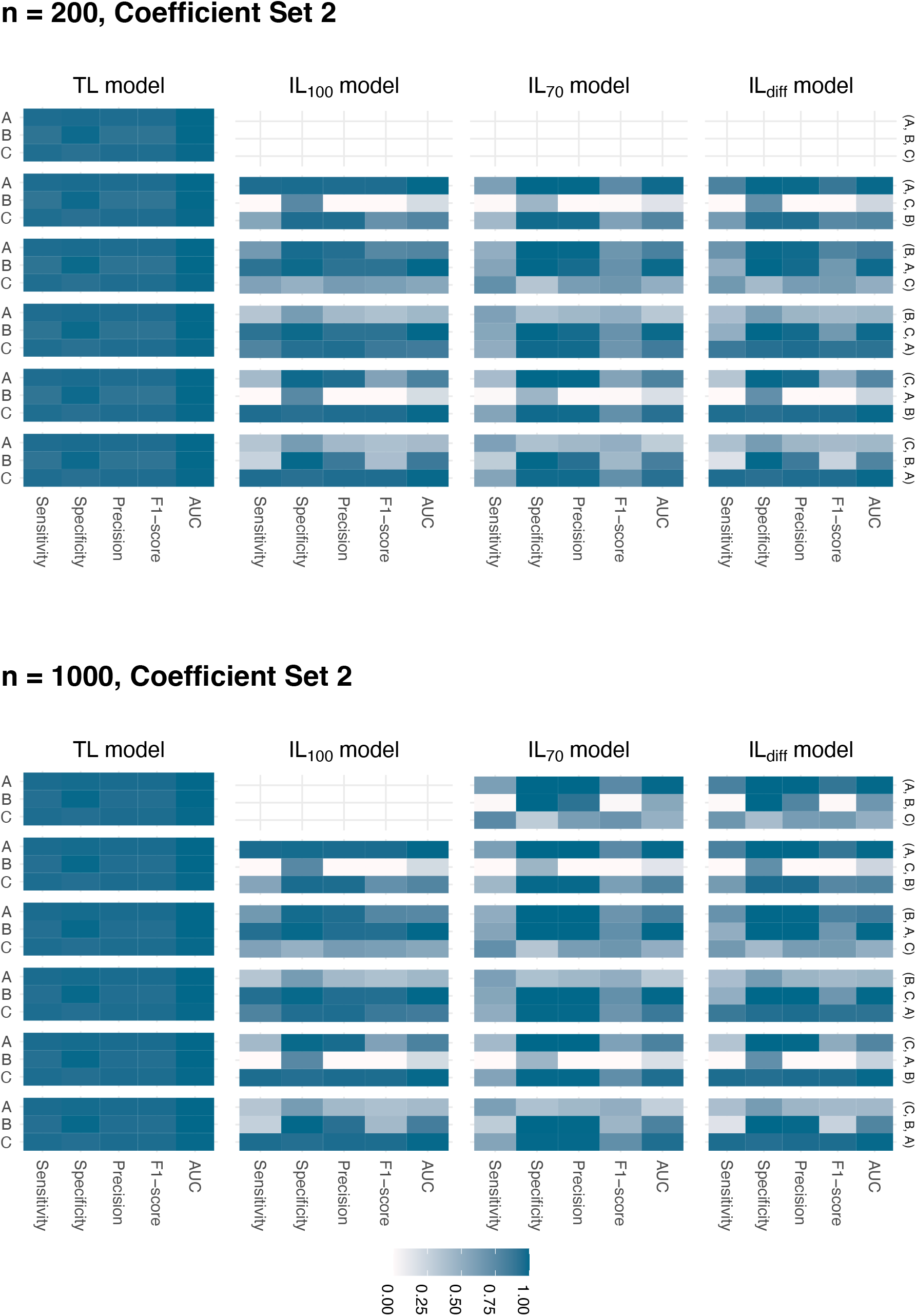
Heatmaps as in Figure 5, here for Coefficient Set 2.

The results show that performance differences between the TL and IL models strongly depend on which of the five metrics is considered: In the overall picture, TL models dominate with respect to AUC and F1-score. They show generally higher sensitivity than the IL_70_ and IL_diff_ models and many IL_100_ models. For specificity and precision, in contrast, the often high performance of the IL_70_ models stands out.

Regarding the individual diseases, the TL model is better in predicting diseases A and C than B; this holds for all performance metrics across all sample sizes, coefficient sets and examination sequences, with the exception of specificity, which is consistently higher for B than for A and C. For the IL models, this pattern is only partly observable. Their prediction performance for individual labels rather varies with the underlying examination sequence: For all IL models, specificity and precision are highest for the first and second disease of each examination sequence. The other way round, the last disease in the sequence is generally predicted worse than the other two; with only few exceptions, the latter observation holds for all IL models, performance metrics, sample sizes and coefficient sets. Moreover, for diseases A and C, it seems to matter which one is tested first, leading to better prediction performance for the earlier one.

For the IL_70_ models, it is striking that the sensitivity and the F1-score are comparably low. This applies to all diseases. The same is observed for the IL_diff_ models; here, the difference in sensitivity and F1-score between the single diseases is more pronounced, in particular for Coefficient Set 1. Overall, the examination sequence clearly affects the predictive ability of the models trained on inaccurately labeled data. The TL models seem to be unaffected by the order of the diseases. Heatmap patterns agree for the considered sample sizes, with occasional differences.

While the previous description of results holds for both coefficient sets, a direct comparison between Figures 5 and 6 reveals that generally better predictive performance is achieved with Coefficient Set 2. In particular, the TL models do not only show clear superiority over all IL models, but also higher reliability for predicting disease B. The latter also applies for the IL models with respect to individual examination sequences: Prediction performance improves especially for those cases where disease B is investigated first.

### 3.3 Discussion

The results of the simulation study clearly demonstrate the superiority of the TL models over all other models independent of the examination sequence, the sample size and the covariate effects. The inferiority of models trained on inaccurately labeled data is only natural as these models are trained on manipulated and tested on non-manipulated data. In particular, the manipulated labelsets are of single-label type and thus do not fully represent the underlying ground truth. In particular, five of the eight possible labelsets are missing in the training data, namely those containing fewer or more than one positive label. The discrepancy between training and test population naturally leads to a large number of incorrectly predicted labelsets. This is penalized particularly strictly by EMR, which only counts completely correctly predicted labelsets, and this explains the pronounced difference in predictive performance observed for this metric.

Regarding the prediction of individual diseases, we primarily observed a decrease in sensitivity and F1-score for the IL models compared to the TL models. This is hardly surprising, particularly for IL_70_ and IL_diff_, since disease occurrences were artificially removed under the assumption of limited diagnostic test-sensitivity; accordingly, these diseases are less likely to be predicted.

While prediction performance of TL models is unaffected by the examination sequence, this picture differs for IL models, where the order strongly affects the reported measures, depending on the position of a disease in the sequence. A direct explanation for this is that our data manipulation process, described in Section 2.3.2, distorts the first test result in a sequence with a lower probability than the second or third: For the second and third tests, the manipulation mimicking limited diagnostic test-sensitivity is compounded by additional manipulation due to misinterpretation. Therefore, it is generally advantageous for a disease to appear early in the sequence.

We observed that disease B was predicted less reliably than A and C, especially regarding sensitivity and F1-score, but with high specificity. We attribute this to two possible reasons. First, the comparably low prevalence of disease B: For the synthetic data generation, we targeted at disease prevalences of 45 % for A, 15 % for B and 60 % for C. Accordingly, the fictional patients suffering from B are seldom, and this can result in pronounced class imbalance. Such class imbalance, in turn, can lead to the minority class being neglected during model estimation (Guo et al., 2008), thereby affecting the prediction. Second, we relate the low prediction performance for disease B to its test-sensitivity in combination with the association to other diseases: The low diagnostic test-sensitivity of 65 % for disease B might have introduced more uncertainty into the target variables, particularly with regard to the association between disease occurrences. As an example, consider a patient who suffers exclusively from disease B and who is first examined for exactly this disease. However, the test result is negative due to low test-sensitivity. Next, the patient is tested for disease A, which is strongly associated to B in a way that the absence of B suggests the presence of A. Consequently, the patient is diagnosed with disease A. In our simulation study, the obtained empirical association between occurrences of the three diseases differs for the two coefficient sets; A and C are hardly associated in both cases, while A and B are moderately associated. For B and C, the association varies between the two sets. Altogether, an incorrect prediction of one disease can come along with an incorrect prediction for the comorbidities.

We investigated the impact of the sample size on the predictive performance of MLC. According to our results, *n* = 200 was sufficiently large such that a further increase of *n* left the performance of the models mostly unaltered. However, a small sample size in combination with a low disease prevalence and low diagnostic test-sensitivity can lead to a complete absence of one class from the generated set of target variables. This becomes particularly relevant if CV is used for model optimization and the size of the set used within one fold of CV is rather small. Consequently, a statistical classification model can maybe not be estimated. This case occurred in our simulation study, particularly for *n* = 200. Even if both classes are present, a low prevalence leads to high class imbalance. In case of a small dataset, reliable methods for class rebalancing can hardly be found. Moreover, small datasets harbor the risk of non-representativeness which can result in model overfitting such that an estimated model is inappropriate for predicting new data. Thus, uncertainty of statistical models decreases with increased sample size; at the same time, computational costs increase with increased sample size.

We have investigated the influence of covariate effect sizes on the models’ performance by generating synthetic data using two exemplary coefficient sets. A main finding is the worse performance for predicting disease B when considering Coefficient Set 1. This set was defined such that the effect of covariates is not restricted to a single disease, but each covariate affects each disease occurrence to some extent. We see a possible explanation for the differing performance in the simultaneous association of disease B with A *and* C, while A and C are hardly associated (see upper part of Figure A3). Accordingly, the presence of disease A, say, may speak against B while not affecting the occurrence of C, but a simultaneous presence of C may speak in favor of B at the same time. This is different for Coefficient Set 2, where A and B are more associated than for Coefficient Set 1, but C is hardly associated with any disease. Accordingly, this selection of coefficients and the resulting disease association can be a reason for B being predicted with higher reliability.

All in all, the results of our simulation study reveal that inaccurately labeled data strongly affects the predictive ability of MLC models. In particular, uncertainty within the diagnostic process can have a massive impact on prediction. Further, the examination sequence influences the models’ performance. We further found out that an increase from 200 to 1000 patients did not substantially affect prediction performance. In contrast, the effect strength of covariates altered the association between disease occurrences and the predictive performance of MLC. We conclude that the sample size of a dataset does not necessarily have to be extensively large, but it is advantageous if covariates are selected specifically for individual diseases.

## 4 Case Study

The simulation study in the previous section has clearly shown that inaccurate labeling decreases the ability of MLC to recognize diseases. This is observed across both investigated sample sizes but depends on the effect strength of covariates as well as on the examination sequence. In this section, we conduct a case study based on real data of chronic pain patients in order to investigate the predictive performance of MLC under more realistic circumstances. We first describe the preprocessing of the dataset, followed by the presentation and discussion the result.

### 4.1 Data

The real dataset, which has already been introduced in Section 2.2, initially contained 246 patients. To create a more homogeneous patient cohort, we exclude 37 patients with previous diagnosis of FMS and other chronic pain syndromes. Afterwards, we reduce the number of covariates from 152 to 21 as follows: We remove covariates collected at primary care because of inconsistencies with measurements collected at secondary care. In addition, we exclude the information from questionnaires to overall scores rather than individual questions since the scores prove more informative. We also remove categorical covariates with a large number of categories as in practice, such covariates are often unsuitable for reliable prediction. Next, we remove covariates which were redundant because their information content is already expressed through other covariates, for example, the body mass index. We also remove dates and covariates which depend on other covariates, for example, the duration of smoking. Finally, we remove covariates with fractions of missing values above 10 % (see Figure A2 in the Appendix).

The remaining dataset still contains missing values. Since health records are highly individual, we aim to restrain from data imputation. As a consequence, we decide to exclusively use complete cases for our analysis, resulting in a dataset of 164 patients. To avoid numerical instability in model estimation, we exclude covariates which cause high correlation or a generalized variance inflation factor larger than five. The covariates remaining in the final dataset are listed and described in Table 4.

**Table 4:**
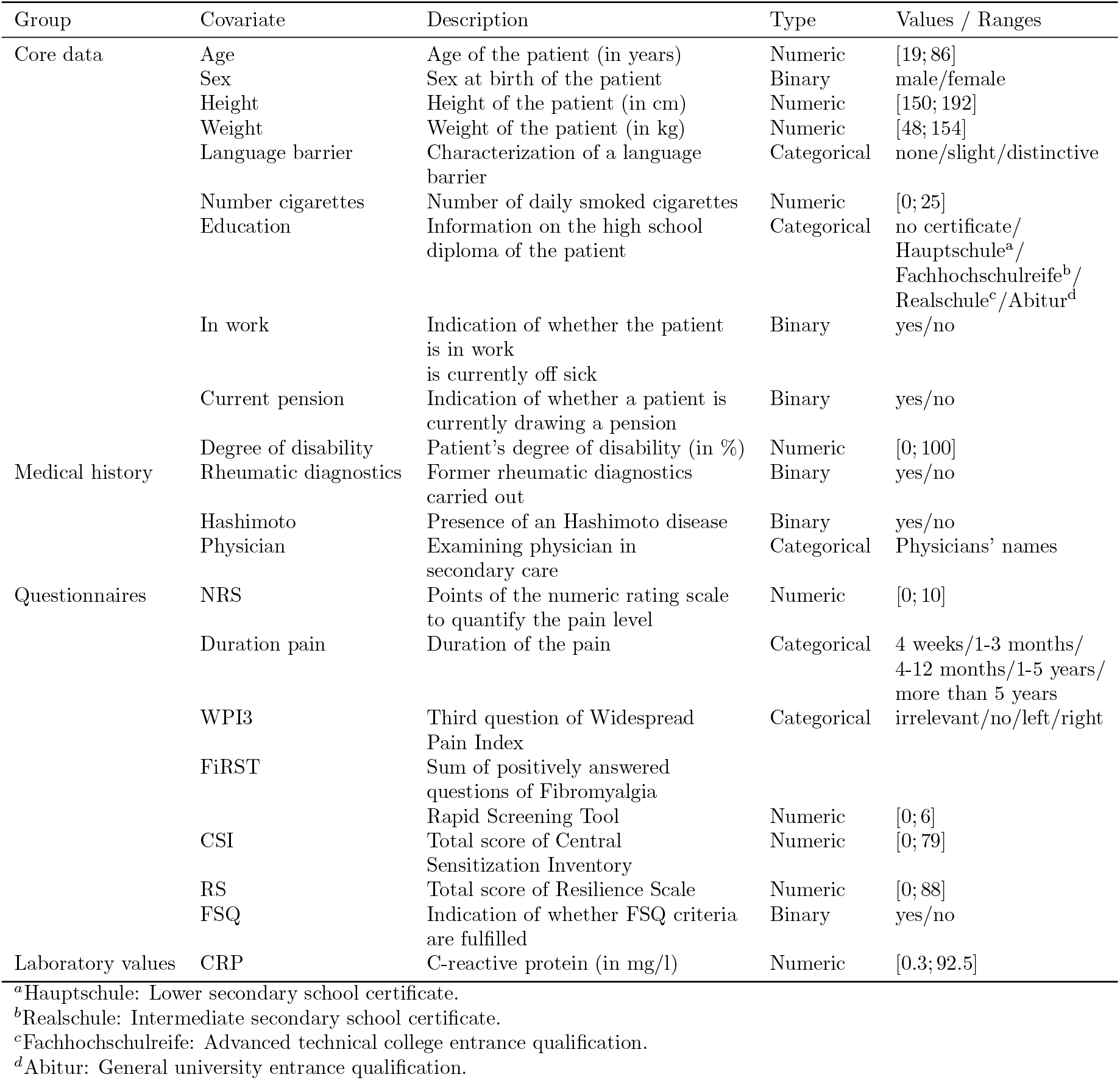
Description of the covariates contained in the final real dataset.

We perform a training-test-split on the final dataset, resulting in a training set containing 80 % (131 patients) of all samples and a test set containing 20 % (33 patients) of the samples. For the training set, we manipulate the target variables according to the procedure described in Section 2.3.2, that means, we derive inaccurately labeled data IL_100_, IL_70_ and IL_diff_ for each possible examination sequence. The test set remains unaltered.

### 4.2 Results

As in the simulation study, we train MLC models on both accurately labeled data as well as on inaccurately labeled data. As label ordering for the CC, we employ the same examination sequences that were used for the simulation of the inaccurately labeled data.

#### Prediction of Full Labelsets

Figure 7 visualizes the performance measures EMR and Jaccard similarity for the considered models; all of them are evaluated on the same test data. As before, we report the Hamming loss in Figure A6 in the Appendix. In addition, a tabular representation of the results is provided in Table A3.

**Figure 7:**
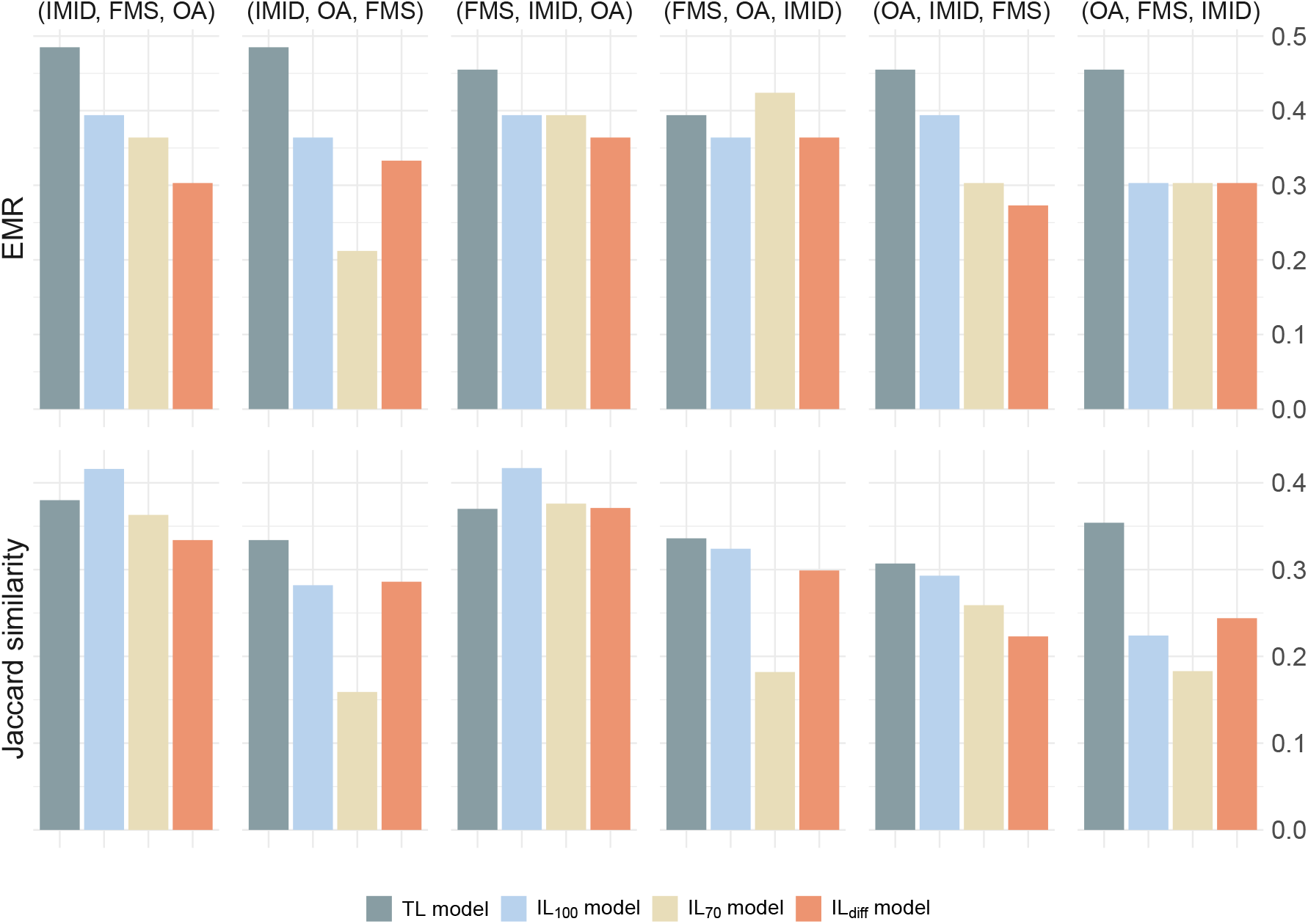
EMR and Jaccard similarity for the models trained on accurately and on inaccurately labeled real data (different colored bars) assuming different examination sequences (groups of four bars each). The model names refer to the employed training datasets. Higher values speak in favor of better model performance. Hamming loss is visualized in Figure A6 in the Appendix. Exact values are listed in Table A3.

TL models achieve highest EMR for all examination sequences except for ‘(FMS, OA, IMID)’, where the IL_70_ model performs slightly better. Overall, EMR is rather low for all models and all examination sequences in comparison to the results from the simulation study in Section 3, as it never exceeds a value of 0.5.

Regarding the less strict Jaccard similarity, TL models perform best for only four out of six examination sequences: For ‘(IMID, FMS, OA)’ and ‘(FMS, IMID, OA)’, one or more IL models show better performance. As for EMR, none of the TL or IL models achieves remarkably good results as compared to those from the simulation study: In the case study, the overall highest value is 0.417, achieved by the IL_100_ model for the sequence ‘(FMS, IMID, OA)’. The picture for Hamming loss is similar.

#### Prediction of Individual Labels

To assess the models’ ability to predict diseases individually, we again consider sensitivity, specificity, precision, F1-score and AUC. Figure 8 visualizes these performance measures for each disease, each model and each examination sequence. Tabular representations of the results are provided in Section A.3 in the Appendix.

**Figure 8:**
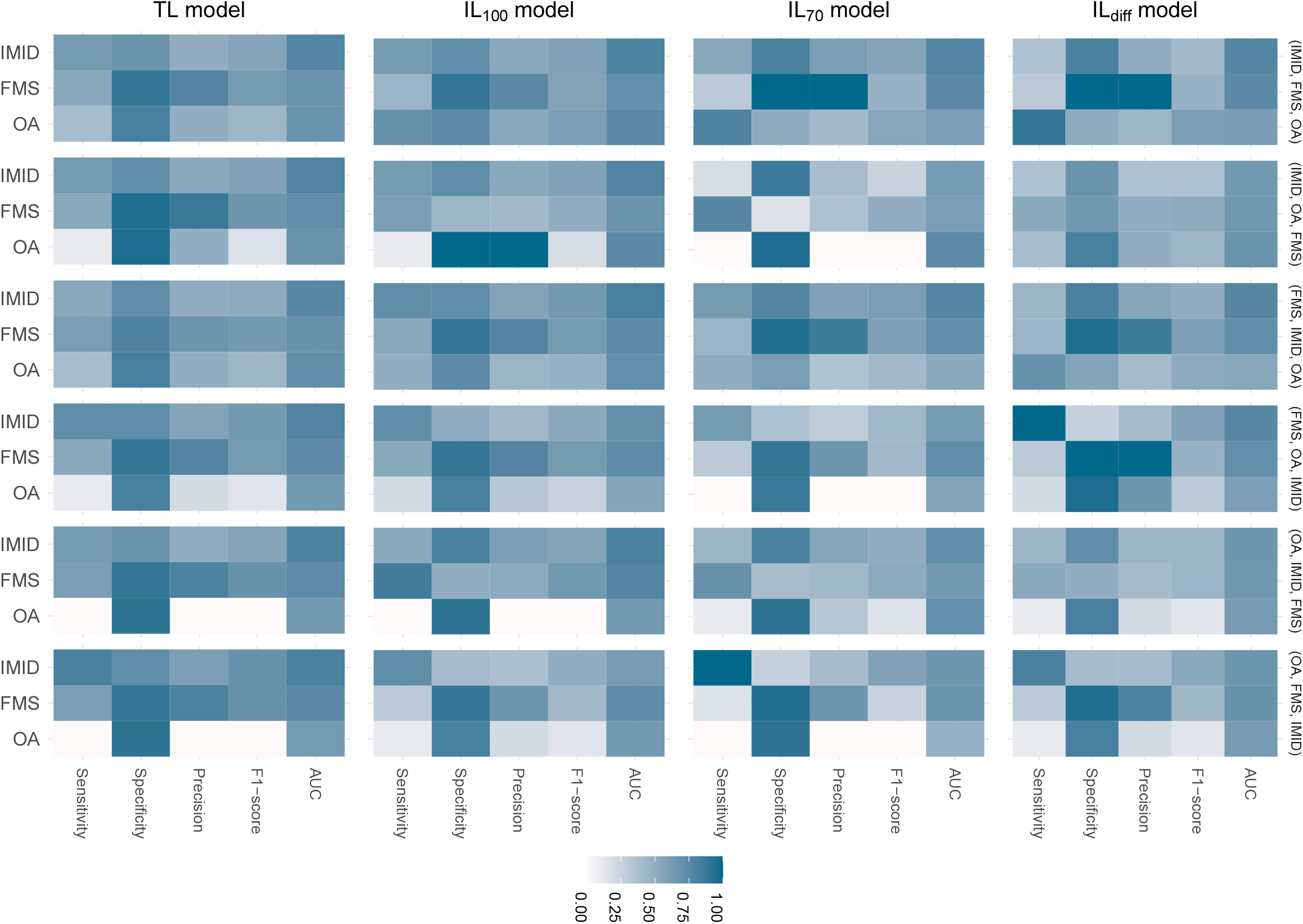
Heatmaps of performance of predicting individual diseases IMID, FMS and OA (see labels on the left) for all models (labels at the top) and examination sequences (labels on the right). Dark colors depict larger values and vice versa. The model names refer to the employed training datasets.

Looking at Figure 8, two observations stand out: First, high values of specificity and precision are repeatedly observed, whereas sensitivity and F1-score tend to be low (with few exceptions for sensitivity); AUC falls within the mid-range. Second, sensitivity, precision and F1-score for OA are most often lower than for IMID and FMS. Specificity and precision for IMID are lower than for FMS unless FMS is the last disease in the examination sequence. On the other hand, the sensitivity for IMID rises when tested last; in those cases, the sensitivity and F1-score for IMID are higher than for the other diseases.

The dominance of TL models with respect to predicting full labelsets becomes less apparent on the level of individual labels: For the examination sequence ‘(OA, FMS, IMID)’, for example, EMR and Jaccard similarity of the TL models lie clearly above those of the IL models. The heatmaps for that sequence, in contrast, show more heterogeneous pictures.

### 4.3 Discussion

In this case study, we applied MLC to accurately and inaccurately labeled real data of chronic pain patients. The results present an ambiguous picture and do not reveal a clear superiority of one model over the others.

We have already discussed in the context of the simulation study that our artificially introduced data manipulation puts the IL models at a disadvantage, as the training and test data follow different data-generating processes. Against this background, a superior performance of the TL models is to be expected, particularly for those metrics that evaluate entire labelsets (EMR, Jaccard similarity, and Hamming loss), and especially for the strict measure EMR. That said, it is striking that this expected advantage of the TL models is not consistently reflected in the performance measures, most notably not in the prediction of individual diseases.

The most remarkable observation is that the examination sequence influences the model performance, particularly if the test-sensitivity is low. In the following, we discuss possible reasons for the ambiguous picture that arises from the case study, particularly taking the results of the simulation study into account. The latter is comparable to the case study regarding the number of diseases, the covariate effect strength, disease association structure and sample size *n* = 200.

#### Covariates

The three considered diseases IMID, FMS and OA can manifest themselves in similar symptoms. However, some covariates are known to be most informative for specific diseases: CRP is particularly relevant for diagnosing IMID (Pope and Choy, 2021), FiRST was specifically developed to identify FMS (Perrot et al., 2010), and X-ray imaging can provide indications of OA (Katz et al., 2021). Our results suggest that a reliable differentiation between these pain-causing diseases using statistical modeling requires the inclusion of informative covariates. Our dataset contains, in particular, specific covariates for diagnosing IMID and FMS, but relevant covariates for OA are missing. This may explain the comparatively low predictive performance of the models for OA. The case study indicates that OA can be better predicted if IMID and FMS are considered before, even for the TL models and (for the IL models) even if the test-sensitivity of the other two diseases is low. The simulation study has shown that predictive performance benefits from covariates that are targeted in the sense that they contribute to explaining specific diseases rather than all of them simultaneously. In contrast, including too many weakly informative or non-specific covariates may lead to a blurring of effects and reduced model performance. Such a situation may be present here, particularly with regard to OA. Furthermore, the available real data seems to be suitable for recognizing patients which do *not* suffer from FMS or OA, expressed through the comparably high specificity.

#### Sample Size

The real dataset contains 164 patients, only 131 of which are used for model training. This low number can affect the results in two aspects: First, class imbalance can become more pronounced and decrease the robustness of classification. The simulation study, however, does not indicate a strong effect of the size of the training sample. Second, the test set of the case study contained only 33 patients, while it was 500 patients for the simulation study. Such a small number of samples harbors the risk of not being representative, i.e., the test set may contain patients with unusual values, making it difficult to make a correct prediction using the pattern learned during model training. In a test set of size 33, individual patients carry more weight, meaning that performance is reduced even with just a few incorrect predictions.

#### Real-data characteristics

The simulation study is based on synthetic data, generated under a well-controlled data-generating process, in particular a logit relationship that explicitly incorporates the covariates later used for training and prediction. Such a structured relationship cannot be guaranteed for real-world data. Therefore, the lower level of the performance measures is only natural, despite the use of prospective data and the estimation of TL models. Apart from that, for real-world data, errors and inconsistencies can hardly be avoided. Example are implausible or improper values, duplicates, different scaling or units. Even after careful preprocessing, inadequate data may remain unnoticed and affect a model’s performance.

Combining the findings of the simulation study and the case study, we conclude that special attention should be paid to two aspects of data collection: on the one hand, all competing or potentially co-occurring diseases under consideration should be fully examined, leading to accurately labeled data; on the other hand, covariates should be specific and informative. If these conditions are met, even a small sample size may be sufficient for reliable MLC. This is particularly advantageous in medical contexts which target the prediction of diseases with low prevalence.

## 5 Conclusion

The starting point of our investigation is the assumption that patients should be considered holistically within diagnostic processes and may, in particular, suffer from multiple diseases simultaneously. A positive or negative diagnosis for one disease may carry information relevant to another diagnosis; however, it should neither automatically lead to its exclusion nor to its assumption by default. MLC provides a suitable statistical framework for modeling and prediction in such settings. The reliability of classification, however, depends on the availability of appropriate data. In practice, one must expect that diagnostic tests may be wrong and that missing information may be misinterpreted. In our study, we aim to investigate the extent to which such inaccurate labeling affects the performance of MLC.

To that end, we conducted a simulation study which considered various scenarios regarding data (mis)interpretation, diagnostic test-sensitivity, sample sizes and the covariate effect strengths. This study enabled us to observe the effects of data manipulation in a controlled setting. It was designed to reflect the data characteristics of the subsequent case study, in which we investigate the prediction of co-occurring pain-causing diseases and likewise introduce artificial data manipulation.

The simulation study clearly indicates the superiority of those models trained on accurately labeled data over models trained on inaccurately labeled data. This holds both for the prediction of complete labelsets and for analyses focusing on individual diseases. In addition, the use of specific and informative covariates increases the models’ performance. These findings are confirmed by the case study which, however, leads to a more ambiguous picture without a clear tendency towards one model. In particular, the advantage of models trained and evaluated on accurately labeled data is markedly smaller than would be expected given the extent of data manipulation. Possible explanations are manifold and include the small sample size, a non-representativeness of the test set as well as the absence of sufficiently informative covariates. On the positive side, it can be noted that the data manipulation does not substantially impair predictive performance in the real-data case. This means that even if data is available in a single-label format for practical reasons, but the underlying context is inherently multi-label, the use of MLC can still be worthwhile.

Future data collection efforts intended to serve as the basis for statistical modeling should therefore take into account two key requirements, which constitute our main conclusion: First, all patients must be comprehensively examined with respect to the diseases of interest, ensuring accurate labeling. Second, the selected covariates should be informative for the diseases under consideration. In addition, a closer investigation of the role of disease associations is of particular importance in order to determine the level of association strength necessary to compensate for potential inaccuracies in the labeling of the data.

While our study accounted for different diagnostic test-sensitivities, motivated by the considered use case, future investigation should take into account the role of specificity as a relevant aspect in ruling out diseases. Another aspect that we have not examined due to limitations of our data basis concerns the information that is inherent in the choice of examination sequence. In our study, we considered this sequence to be randomly chosen. In clinical practice, however, this sequence is determined by factors such as urgency, available resources, costs, and the invasiveness of diagnostic procedures. In addition, the physician’s level of suspicion may play a role, so that the condition considered least likely is investigated last, whereas the most probable or life-threatening conditions are addressed first. Given that our results partly indicate a particular role for the disease positioned at the end of the sequence, this aspect warrants closer investigation.

Interpreting our results, we conclude that incomplete diagnoses in the medical field, leading to inaccurately labeled data, can substantially affect the performance of MLC. Consequently, when developing statistical models for disease recognition in settings where the disease of interest is frequently accompanied by comorbidities, particular care must be taken in data collection. Specifically, patients should be comprehensively examined with respect to all diseases under consideration to ensure accurate labeling, and the selected covariates should be targeted and informative for the respective conditions. By accounting for these aspects, our findings may contribute to improved study design and to a more informed and critical interpretation of data used for statistical modeling.

## Data Availability

The data of the German hospital cannot be shared due to data privacy protection.

## Glossary

AI: artificial intelligence
AUC: area under the curve
CC: classifier chain
CDSS: clinical decision support system
CRP: C-reactive protein
CSI: Central Sensitization Inventor
CV: cross-validation
EMR: exact match ratio
FiRST: Fibromyalgia Rapid Screening Tool
FMS: fibromyalgia syndrome
IMID: immune-mediated inflammatory diseases
LRM: logistic regression model
MLC: multi-label classification
MLC: multi-label classification
NRS: numeric rating scale
MLC: multi-label classification
OA: osteoarthritis
MLC: multi-label classification
SLC: single-label classification
MLC: multi-label classification
WPI: Widespread Pain Index

## Author Contributions

Conceptualization: Sophie Schmiegel, Christiane Fuchs

Data curation: Sophie Schmiegel, Martin Rudwaleit, Marvin-Hendrik Röchter

Formal analysis: Sophie Schmiegel

Funding acquisition: Christiane Fuchs, Martin Rudwaleit

Methodology: Sophie Schmiegel, Christiane Fuchs

Supervision: Christiane Fuchs

Visualization: Sophie Schmiegel

Writing – original draft: Sophie Schmiegel

Writing – review and editing: Sophie Schmiegel, Christiane Fuchs, Hannah Marchi, Martin Rudwaleit, Marvin-Hendrik Röchter

## Conflict of interest

None declared.

## Funding

This work was supported by the Medical Research Start-up Fund of the Medical School OWL, Bielefeld University.

## Ethics Statement and Consent to Participate

Ethical approval was given by the Ethics Committee of the Westphalia-Lippe Medical Association and the University of Münster (Ethik-Kommission der Ä rztekammer Westfalen-Lippe und der Westfälischen Wilhelms-Universität Münster); case number: 2021-426-f-S. All patients agreed on participating in this study.

## Computational Details and Data Availability Statement

The data preprocessing and the synthetic data generation were performed using R version 4.5.2 (R Core Team, 2025). Data manipulation was done using R version 4.6.1 (R Core Team, 2026). The models were estimated using Python version 3.12.13 (Python Software Foundation, 2026).

The code used to generate the synthetic data, for data manipulation and for model estimation is available at https://github.com/fuchslab/Inaccurately_Labeled_Data. The data of the German hospital cannot be shared due to data privacy protection.

## Acknowledgements

We thank Tamara Schamberger and Valentina Lakusta for helpful comments and stimulating discussions.

## APPENDIX A

### A.1 Additional Material on ‘Clinical Use Case and Real Data Basis’

**Figure A1:**
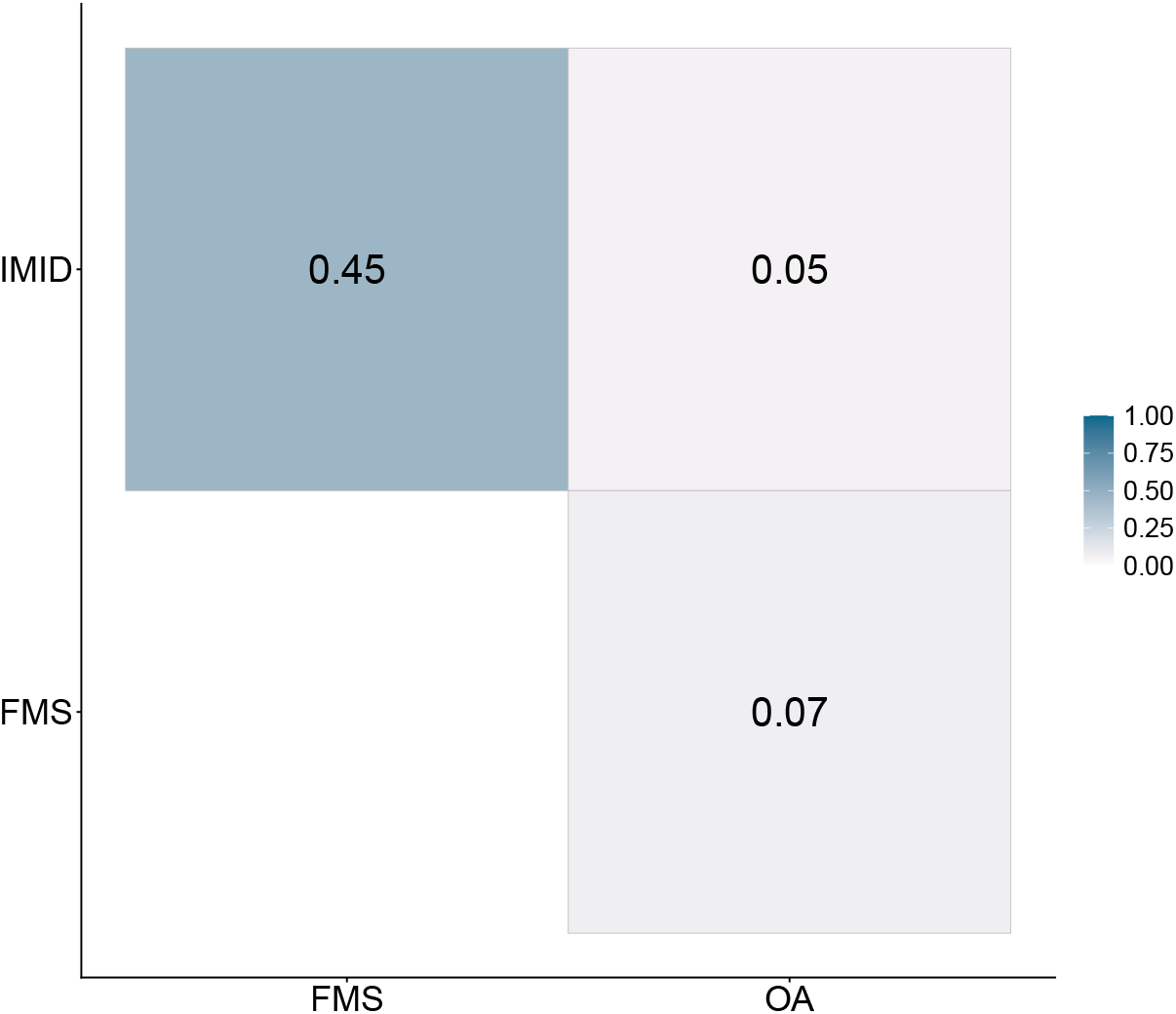
Cramér’s V for the three considered diseases IMID, FMS and OA in the final real dataset.

**Figure A2:**
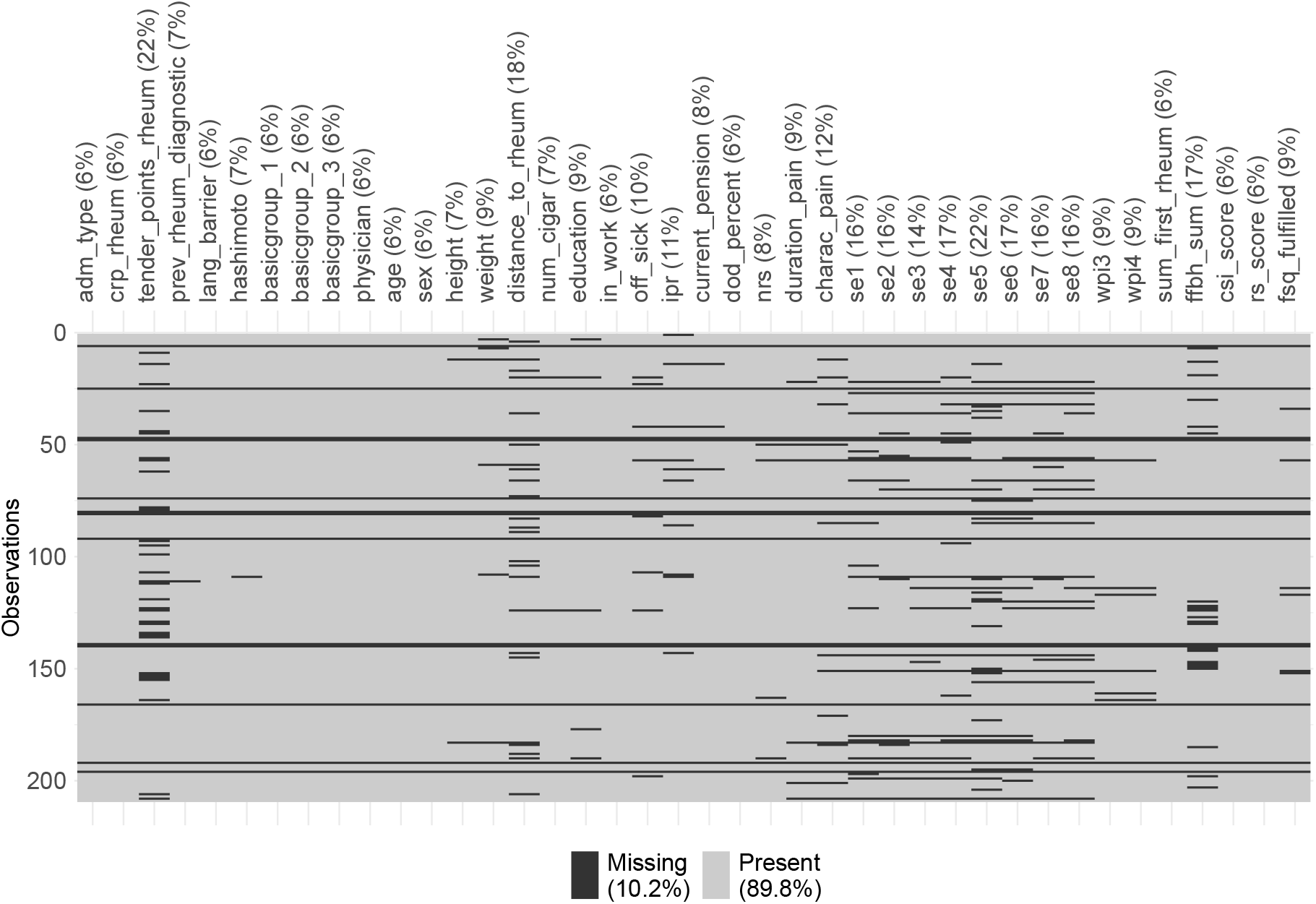
Frequencies of missing values in the real data after preprocessing as described in Section 4.1. The heatmap indicates missing and present values per sample (rows, arbitrary order) and covariate (columns). The further removal of covariates with frequencies of missing values above 10 % and the subsequent exclusion of incomplete cases results in the final dataset containing 24 covariates of 164 patients.

**Table A1:**
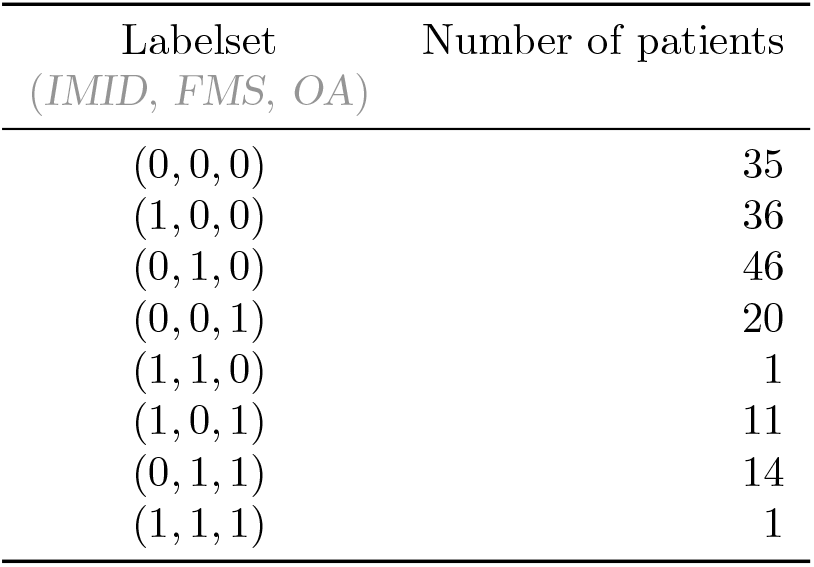
Number of occurrences for each labelset among the 164 patients in the final real dataset.

| Labelset<br>(*IMID*, *FMS*, *OA*) | Number of patients |
| --- | --- |
| (0, 0, 0) | 35 |
| (1, 0, 0) | 36 |
| (0, 1, 0) | 46 |
| (0, 0, 1) | 20 |
| (1, 1, 0) | 1 |
| (1, 0, 1) | 11 |
| (0, 1, 1) | 14 |
| (1, 1, 1) | 1 |

### A.2 Additional Material on the Simulation Study

**Figure A3:**
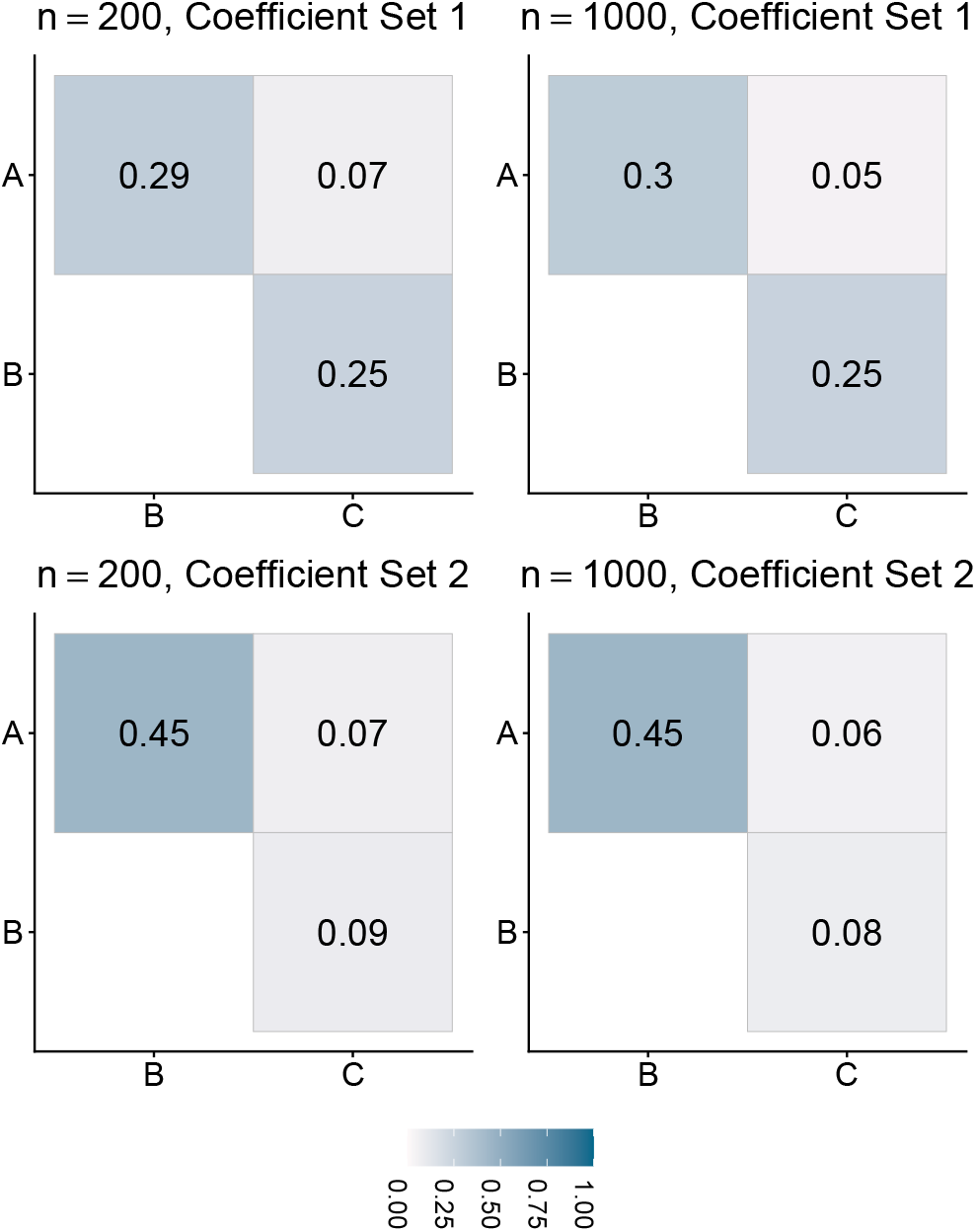
Cramér’s V for simulated occurrences of diseases A, B and C according to TL training sets, averaged across 500 replicates for *n* = 200 as well as *n* = 1000 and both coefficient sets.

**Figure A4:**
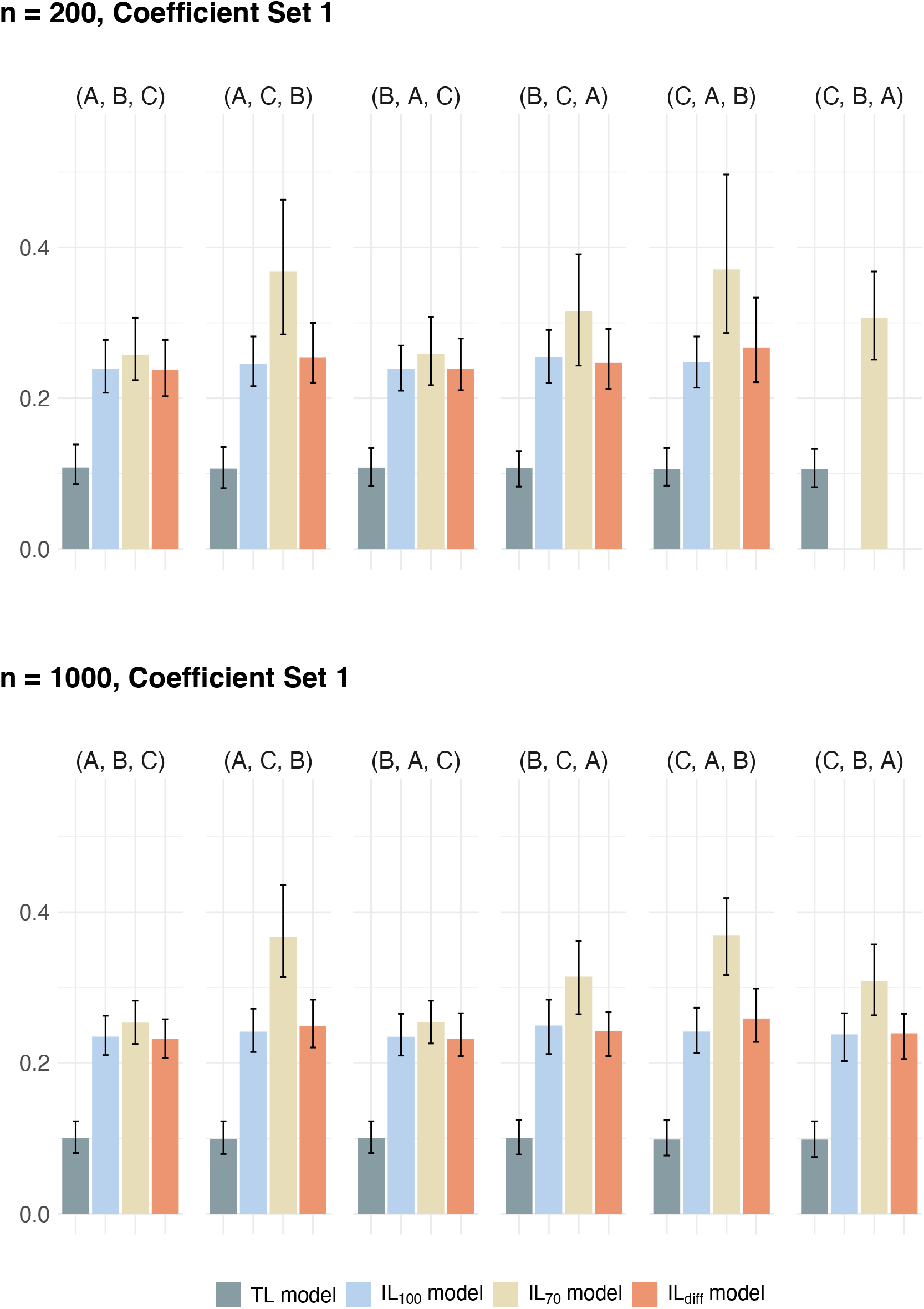
Hamming loss for the models trained on accurately and on inaccurately labeled synthetic data (different colored bars) considering Coefficient Set 1 for the data generation process and assuming different examination sequences (groups of four bars each). The model names refer to the employed training datasets. Lower values speak in favor of better model performance. Corresponding EMR and Jaccard similarity are visualized in Figure 3 in the main text.

**Figure A5:**
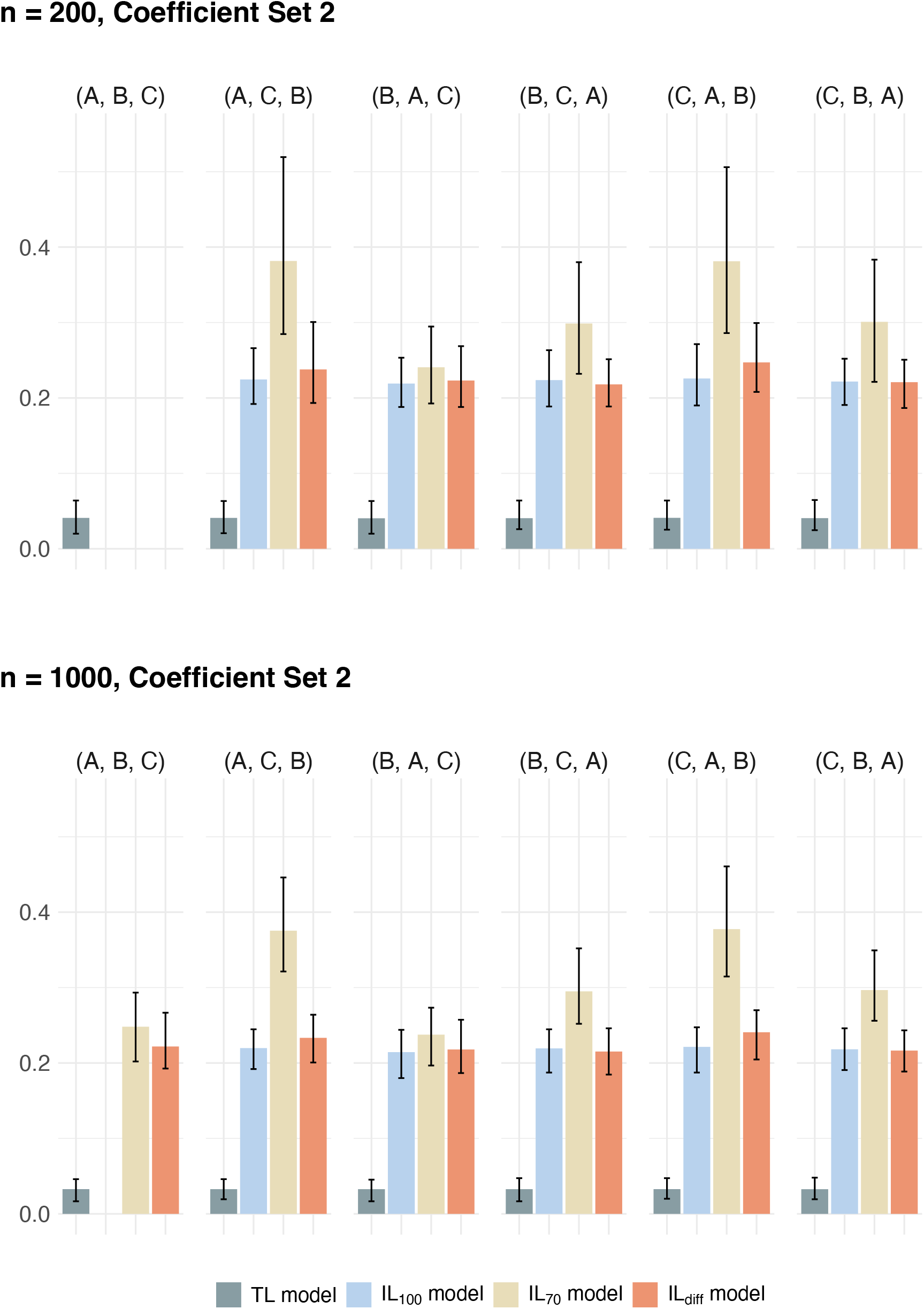
Hamming loss as in Figure A4, here for Coefficient Set 2. Corresponding EMR and Jaccard similarity are visualized in Figure 4 in the main text.

**Table A2:** Regression coefficients for synthetic data generation for the simulation study.

| | Disease | $\beta_0$ | $\beta_1$ | $\beta_2$ | $\beta_3$ | $\beta_4$ | $\beta_5$ | $\beta_6$ |
| --- | --- | --- | --- | --- | --- | --- | --- | --- |
| Coefficient Set 1 | A | -0.386 | 0.207 | -0.797 | 0.668 | -0.401 | 0.407 | -0.236 |
|  | B | -0.948 | -0.265 | 0.204 | -0.390 | 1.215 | 0.239 | 0.284 |
|  | C | -1.035 | 0.754 | -0.021 | -0.284 | 0.150 | -0.109 | -0.047 |
| Coefficient Set 2 | A | -0.4 | 0.0 | -0.8 | 3.4 | 0.0 | 0.0 | -1.8 |
|  | B | -1.0 | 1.1 | -1.2 | 2.5 | -3.2 | 1.6 | -2.8 |
|  | C | -1.0 | 2.1 | 0.0 | 0.0 | -2.0 | 1.3 | 0.0 |

### A.3 Additional Material on the Case Study

**Figure A6:**
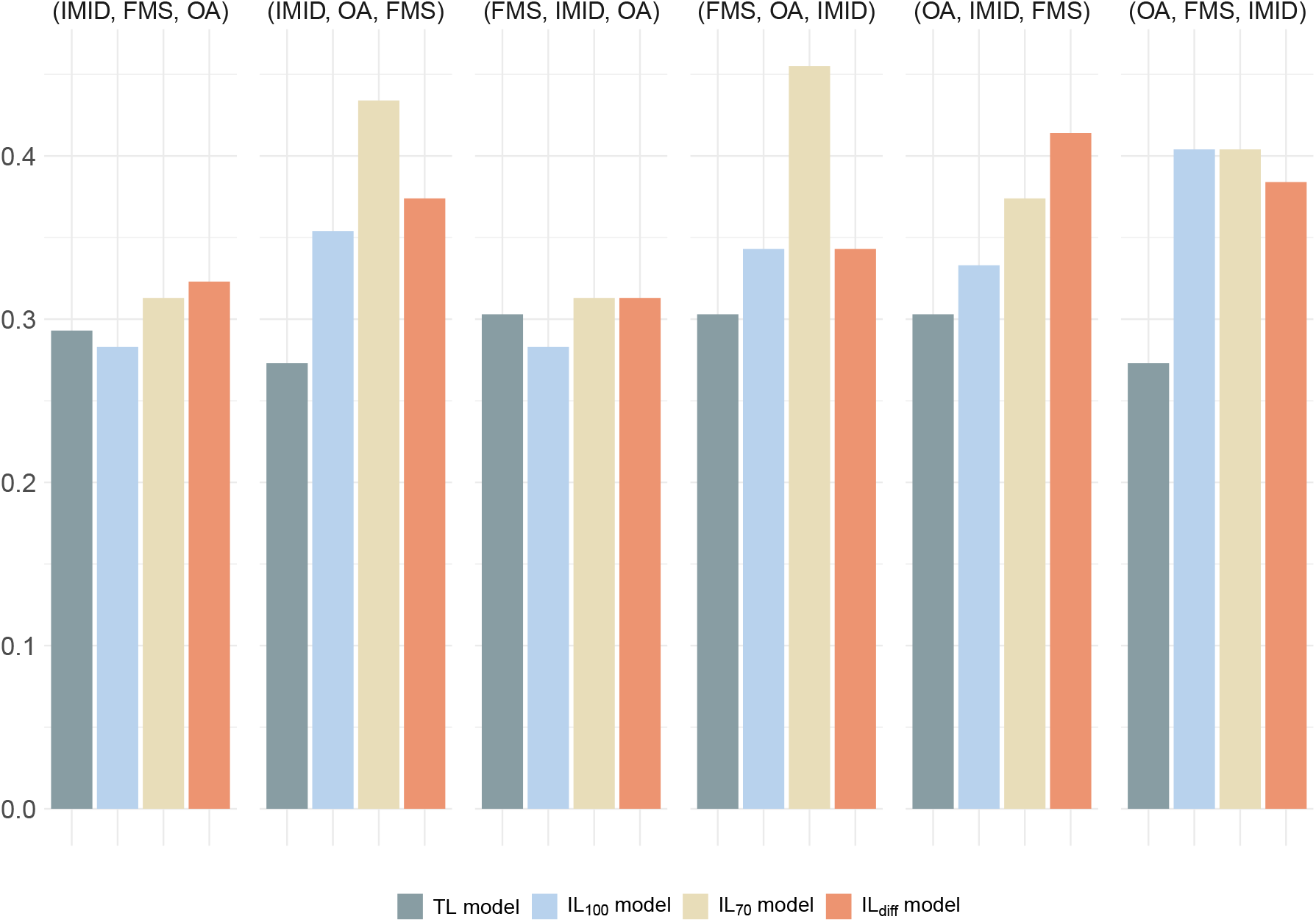
Hamming loss for the models trained on accurately and on inaccurately labeled real data (different colored bars) assuming different examination sequences (groups of four bars each). The model names refer to the employed training datasets. Lower values speak in favor of better model performance. Corresponding EMR and Jaccard similarity are visualized in Figure 7 in the main text.

**Table A3:** Multi-label performance measures obtained in the case study. MLC models are trained on accurately and on inaccurately labeled data, assuming various diagnostic examination sequences. The results are also visualized in Figures 7 and A6.

| Examination sequence | Training data | Test-Sensitivity | EMR | Jaccard similarity | Hamming loss |
| --- | --- | --- | --- | --- | --- |
| (IMID, FMS, OA) | TL | – | 0.485 | 0.380 | 0.293 |
|  | IL<sub>100</sub> | 100 % | 0.394 | 0.416 | 0.283 |
|  | IL<sub>70</sub> | 70 % | 0.364 | 0.364 | 0.313 |
|  | IL<sub>diff</sub> | 85 / 65 / 100 % | 0.303 | 0.334 | 0.323 |
| (IMID, OA, FMS) | TL | – | 0.485 | 0.334 | 0.273 |
|  | IL<sub>100</sub> | 100 % | 0.364 | 0.282 | 0.354 |
|  | IL<sub>70</sub> | 70 % | 0.212 | 0.159 | 0.434 |
|  | IL<sub>diff</sub> | 85 / 65 / 100 % | 0.333 | 0.286 | 0.374 |
| (FMS, IMID, OA) | TL | – | 0.455 | 0.370 | 0.303 |
|  | IL<sub>100</sub> | 100 % | 0.394 | 0.417 | 0.283 |
|  | IL<sub>70</sub> | 70 % | 0.394 | 0.376 | 0.313 |
|  | IL<sub>diff</sub> | 85 / 65 / 100 % | 0.364 | 0.371 | 0.313 |
| (FMS, OA, IMID) | TL | – | 0.394 | 0.336 | 0.303 |
|  | IL<sub>100</sub> | 100 % | 0.364 | 0.324 | 0.343 |
|  | IL<sub>70</sub> | 70 % | 0.455 | 0.182 | 0.242 |
|  | IL<sub>diff</sub> | 85 / 65 / 100 % | 0.364 | 0.299 | 0.343 |
| (OA, IMID, FMS) | TL | – | 0.455 | 0.307 | 0.303 |
|  | IL<sub>100</sub> | 100 % | 0.394 | 0.293 | 0.333 |
|  | IL<sub>70</sub> | 70 % | 0.303 | 0.259 | 0.374 |
|  | IL<sub>diff</sub> | 85 / 65 / 100 % | 0.273 | 0.223 | 0.414 |
| (OA, FMS, IMID) | TL | – | 0.455 | 0.354 | 0.273 |
|  | IL<sub>100</sub> | 100 % | 0.303 | 0.224 | 0.404 |
|  | IL<sub>70</sub> | 70 % | 0.303 | 0.183 | 0.404 |
|  | IL<sub>diff</sub> | 85 / 65 / 100 % | 0.303 | 0.244 | 0.384 |

**Table A4:** Performance measures obtained in the case study for each individual disease obtained by MLC, here for the examination sequence ‘(IMID, FMS, OA)’. These values are also visualized in Figure 8.

| Metric | *TL model* |  |  | *IL<sub>100</sub> model* |  |  | *IL<sub>70</sub> model* |  |  | *IL<sub>diff</sub> model* |  |  |
| --- | --- | --- | --- | --- | --- | --- | --- | --- | --- | --- | --- | --- |
|  | IMID | FMS | OA | IMID | FMS | OA | IMID | FMS | OA | IMID | FMS | OA |
| Sensitivity | 0.636 | 0.538 | 0.400 | 0.636 | 0.462 | 0.700 | 0.545 | 0.308 | 0.800 | 0.364 | 0.308 | 0.900 |
| Specificity | 0.682 | 0.900 | 0.826 | 0.727 | 0.900 | 0.739 | 0.818 | 1.000 | 0.522 | 0.818 | 1.000 | 0.522 |
| Precision | 0.500 | 0.778 | 0.500 | 0.538 | 0.750 | 0.538 | 0.600 | 1.000 | 0.421 | 0.500 | 1.000 | 0.450 |
| F1-score | 0.560 | 0.636 | 0.444 | 0.583 | 0.571 | 0.609 | 0.571 | 0.471 | 0.552 | 0.421 | 0.471 | 0.600 |
| AUC | 0.777 | 0.688 | 0.687 | 0.798 | 0.712 | 0.752 | 0.789 | 0.746 | 0.596 | 0.777 | 0.746 | 0.617 |

**Table A5:** Performance measures as in Table A4, here for examination sequence ‘(IMID, OA, FMS)’.

| Metric | *TL model* |  |  | *IL<sub>100</sub> model* |  |  | *IL<sub>70</sub> model* |  |  | *IL<sub>diff</sub> model* |  |  |
| --- | --- | --- | --- | --- | --- | --- | --- | --- | --- | --- | --- | --- |
|  | IMID | FMS | OA | IMID | FMS | OA | IMID | FMS | OA | IMID | FMS | OA |
| Sensitivity | 0.636 | 0.538 | 0.100 | 0.636 | 0.615 | 0.100 | 0.182 | 0.769 | 0.000 | 0.364 | 0.538 | 0.400 |
| Specificity | 0.727 | 0.950 | 0.957 | 0.727 | 0.450 | 1.000 | 0.864 | 0.150 | 0.957 | 0.682 | 0.650 | 0.826 |
| Precision | 0.538 | 0.875 | 0.500 | 0.538 | 0.421 | 1.000 | 0.400 | 0.370 | 0.000 | 0.364 | 0.500 | 0.500 |
| F1-score | 0.583 | 0.667 | 0.167 | 0.583 | 0.500 | 0.182 | 0.250 | 0.500 | 0.000 | 0.364 | 0.519 | 0.444 |
| AUC | 0.793 | 0.727 | 0.687 | 0.793 | 0.681 | 0.748 | 0.632 | 0.615 | 0.743 | 0.649 | 0.650 | 0.683 |

**Table A6:** Performance measures as in Table A4, here for examination sequence ‘(FMS, IMID, OA)’.

| Metric | *TL model* |  |  | *IL<sub>100</sub> model* |  |  | *IL<sub>70</sub> model* |  |  | *IL<sub>diff</sub> model* |  |  |
| --- | --- | --- | --- | --- | --- | --- | --- | --- | --- | --- | --- | --- |
|  | IMID | FMS | OA | IMID | FMS | OA | IMID | FMS | OA | IMID | FMS | OA |
| Sensitivity | 0.545 | 0.615 | 0.400 | 0.727 | 0.538 | 0.500 | 0.636 | 0.462 | 0.500 | 0.455 | 0.462 | 0.700 |
| Specificity | 0.727 | 0.800 | 0.826 | 0.727 | 0.900 | 0.739 | 0.773 | 0.950 | 0.609 | 0.818 | 0.950 | 0.565 |
| Precision | 0.500 | 0.667 | 0.500 | 0.571 | 0.778 | 0.455 | 0.583 | 0.857 | 0.357 | 0.556 | 0.857 | 0.412 |
| F1-score | 0.522 | 0.640 | 0.444 | 0.640 | 0.636 | 0.476 | 0.609 | 0.600 | 0.417 | 0.500 | 0.600 | 0.519 |
| AUC | 0.764 | 0.692 | 0.717 | 0.826 | 0.738 | 0.713 | 0.777 | 0.727 | 0.535 | 0.769 | 0.727 | 0.548 |

**Table A7:** Performance measures as in Table A4, here for examination sequence ‘(FMS, OA, IMID)’.

| Metric | *TL model* |  |  | *IL<sub>100</sub> model* |  |  | *IL<sub>70</sub> model* |  |  | *IL<sub>diff</sub> model* |  |  |
| --- | --- | --- | --- | --- | --- | --- | --- | --- | --- | --- | --- | --- |
|  | IMID | FMS | OA | IMID | FMS | OA | IMID | FMS | OA | IMID | FMS | OA |
| Sensitivity | 0.727 | 0.538 | 0.100 | 0.727 | 0.538 | 0.200 | 0.636 | 0.308 | 0.000 | 1.000 | 0.308 | 0.200 |
| Specificity | 0.727 | 0.900 | 0.826 | 0.500 | 0.900 | 0.826 | 0.364 | 0.900 | 0.739 | 0.273 | 1.000 | 0.957 |
| Precision | 0.571 | 0.778 | 0.200 | 0.421 | 0.778 | 0.333 | 0.333 | 0.667 | 0.000 | 0.407 | 1.000 | 0.667 |
| F1-score | 0.640 | 0.636 | 0.133 | 0.533 | 0.636 | 0.250 | 0.438 | 0.421 | 0.000 | 0.579 | 0.471 | 0.308 |
| AUC | 0.793 | 0.738 | 0.639 | 0.702 | 0.735 | 0.565 | 0.636 | 0.727 | 0.565 | 0.760 | 0.708 | 0.596 |

**Table A8:** Performance measures as in Table A4, here for examination sequence ‘(OA, IMID, FMS)’.

| Metric | *TL model* |  |  | *IL<sub>100</sub> model* |  |  | *IL<sub>70</sub> model* |  |  | *IL<sub>diff</sub> model* |  |  |
| --- | --- | --- | --- | --- | --- | --- | --- | --- | --- | --- | --- | --- |
|  | IMID | FMS | OA | IMID | FMS | OA | IMID | FMS | OA | IMID | FMS | OA |
| Sensitivity | 0.636 | 0.615 | 0.000 | 0.545 | 0.846 | 0.000 | 0.455 | 0.692 | 0.100 | 0.455 | 0.538 | 0.100 |
| Specificity | 0.682 | 0.900 | 0.913 | 0.818 | 0.500 | 0.913 | 0.818 | 0.400 | 0.913 | 0.727 | 0.500 | 0.826 |
| Precision | 0.500 | 0.800 | 0.000 | 0.600 | 0.524 | 0.000 | 0.556 | 0.429 | 0.333 | 0.455 | 0.412 | 0.200 |
| F1-score | 0.560 | 0.696 | 0.000 | 0.571 | 0.647 | 0.000 | 0.500 | 0.529 | 0.154 | 0.455 | 0.467 | 0.133 |
| AUC | 0.806 | 0.735 | 0.643 | 0.814 | 0.781 | 0.643 | 0.719 | 0.638 | 0.713 | 0.657 | 0.650 | 0.630 |

**Table A9:** Performance measures as in Table A4, here for examination sequence ‘(OA, FMS, IMID)’.

| Metric | *TL model* |  |  | *IL<sub>100</sub> model* |  |  | *IL<sub>70</sub> model* |  |  | *IL<sub>diff</sub> model* |  |  |
| --- | --- | --- | --- | --- | --- | --- | --- | --- | --- | --- | --- | --- |
|  | IMID | FMS | OA | IMID | FMS | OA | IMID | FMS | OA | IMID | FMS | OA |
| Sensitivity | 0.818 | 0.615 | 0.000 | 0.727 | 0.308 | 0.100 | 1.000 | 0.154 | 0.000 | 0.818 | 0.308 | 0.100 |
| Specificity | 0.727 | 0.900 | 0.913 | 0.409 | 0.900 | 0.826 | 0.273 | 0.950 | 0.913 | 0.409 | 0.950 | 0.826 |
| Precision | 0.600 | 0.800 | 0.000 | 0.381 | 0.667 | 0.200 | 0.407 | 0.667 | 0.000 | 0.409 | 0.800 | 0.200 |
| F1-score | 0.692 | 0.696 | 0.000 | 0.500 | 0.421 | 0.133 | 0.579 | 0.250 | 0.000 | 0.545 | 0.444 | 0.133 |
| AUC | 0.818 | 0.746 | 0.635 | 0.624 | 0.731 | 0.643 | 0.661 | 0.681 | 0.474 | 0.669 | 0.692 | 0.630 |

## Notes

### Competing Interest Statement

The authors have declared no competing interest.

### Author Declarations

Ethics Committee of the Westphalia-Lippe Medical Association and the University of Muenster (Ethik-Kommission der Aerztekammer Westfalen-Lippe und der Westfaelischen Wilhelms-Universitaet Muenster) gave ethical approval for this work.

